# Post Hoc Evaluation of Probabilistic Model Forecasts: A COVID-19 Case Study

**DOI:** 10.1101/2020.12.09.20246157

**Authors:** Kyle J. Colonna, Roger M. Cooke, John S. Evans

## Abstract

To combat the spread of coronavirus disease 2019 (COVID-19), decision-makers and the public may desire forecasts of the cases, hospitalizations, and deaths that are likely to occur. Thankfully, dozens of COVID-19 forecasting models exist and many of their forecasts have been made publicly available. However, there has been little published peer-reviewed information regarding the performance of these models and what is available has focused mostly on the performance of their central estimates (i.e., predictive performance). There has been little reported on the accuracy of their uncertainty estimates (i.e., probabilistic performance), which could inform users how often they would be surprised by observations outside forecasted confidence intervals. To address this gap in knowledge, we borrow from the literature on formally elicited expert judgment to demonstrate one commonly used approach for resolving this issue. For two distinct periods of the pandemic, we applied the Classical Model (CM) to evaluate probabilistic model performance and constructed a performance-weighted ensemble based on this evaluation. Some models which exhibited good predictive performance were found to have poor probabilistic performance, and vice versa. Only two of the nine models considered exhibited superior predictive and probabilistic performance. Additionally, the CM-weighted ensemble outperformed the equal-weighted and predictive-weighted ensembles. With its limited scope, this study does not provide definitive conclusions on model performance. Rather, it highlights the evaluation methodology and indicates the utility associated with using the CM when assessing probabilistic performance and constructing high performing ensembles, not only for COVID-19 modeling but other applications as well.

**Significance Statement:** Coronavirus disease 2019 (COVID-19) forecasting models can provide critical information for decision-makers and the public. Unfortunately, little information on their performance has been published, particularly regarding the accuracy of their uncertainty estimates (i.e., probabilistic performance). To address this research gap, we demonstrate the Classical Model (CM), a commonly used approach from the literature on formally elicited expert judgment, which considers both the tightness of forecast confidence intervals and frequency in which confidence intervals contain the observation. Two models exhibited superior performance and the CM-based ensemble consistently outperformed the other constructed ensembles. While these results are not definitive, they highlight the evaluation methodology and indicate the value associated with using the CM when assessing probabilistic performance and constructing high performing ensembles.

## 1. Introduction

Effective non-pharmaceutical interventions (NPIs), or community mitigation strategies, are crucial in combating the spread of contagious illnesses like coronavirus disease 2019 (COVID-19) (1). Community NPIs, such as social distancing guidelines, restrictions, closures, and lockdowns, can effectively delay and diminish an epidemic peak (1-3), also known as flattening the epidemic curve (2). However, these NPIs can also have immediate educational and economic consequences (4-6). To make decisions on the implementation of community NPIs amidst the COVID-19 pandemic, the relevant decision-makers (e.g., government officials, community leaders, school administrators etc.) may desire estimates of the number of coronavirus cases, hospitalizations, and deaths likely to occur in their region of interest within the coming weeks.

Information about, and forecasts from, dozens of COVID-19 forecasting models have been made available via the University of Massachusetts Amherst Reich Lab’s COVID-19 Forecast Hub (7). All the modeling groups that have participated have provided predictions (or, central estimates), and most also quantitatively characterize the uncertainty in their predictions – often giving an interquartile range (25% LCL to 75% UCL) and a 90% confidence interval (5% LCL to 95% UCL) for each prediction (8). These models could serve as incredible resources for decision-makers and the public. To understand how much confidence is warranted based on a model’s forecasts, they must have access to evidence about the performance of these models (i.e., how well do forecasts correspond with observations).

Unfortunately, there has been little published peer-reviewed information regarding the performance of COVID-19 forecasting models. However, one study has investigated their predictive accuracy (i.e., how close are predictions to the actual observed values). The authors compared the median absolute percent error (MAPE) of predictions from seven individual models, stratified across world region, month of model estimation, and weeks of extrapolation (9). Collectively, the seven models’ forecasts over a twelve-week range had a MAPE of about 27%, with errors tending to increase with longer forecasts and the best performing model varying by region (9).

While this information is quite useful and provides a sense of the typical bias of model predictions, other aspects of model performance may also be of interest. Users may also care about a model’s precision in its predictions (i.e., how close are point estimates to each other relative to the actual observed values) and its probabilistic performance (i.e., the modeler’s ability to properly characterize the uncertainty in their predictions). A model’s prediction does not fully capture what the modeling group believes might occur over a given time range; they are not claiming that their prediction will occur with 100% likelihood. The probability density provided by a model’s forecast is just as important as their prediction, as the range of this probability density could suggest that a decision-maker takes decisive action for their next step, or exercises caution. Without such information, decision-makers and the public have no sense of how often they will be surprised by observations that are well outside the confidence intervals of predictions provided by the modelers. Frequent surprises may cause the public to lose confidence in the forecasting process and render a potentially valuable tool in the arsenal of COVID-19 mitigation strategies effectively useless.

Scientists concerned with the use of formally elicited expert judgment in support of public policy have faced this issue for decades and have devised approaches for evaluating the probabilistic performance of expert judgments. In this case, models may serve as the ‘experts’ and their forecasts may serve as their ‘judgments’. One such approach, that can evaluate the probabilistic performance of judgments post hoc, is the Classical Model (CM) method (10). This approach has been employed in many studies (11-13) and has been suggested/implemented in other studies concerned with model forecasting (14, 15). Forty-nine applications since 2006 are referenced and analyzed in the recent Cooke, Marti, & Mazzuchi (2021) study (16). Other protocols for formally elicited expert judgment studies (e.g., IDEA [investigate, discuss, estimate, and aggregate] and the Sheffield elicitation framework [SHELF]), require behavioral aggregation or plenary expert deliberation which are unsuitable for post hoc analysis (17, 18).

Some may also wonder whether better forecasts might be obtained by averaging the forecasts of two or more individual models. The Reich Lab has created one such ensemble model (19), which has been prominently featured by the Centers for Disease Control and Prevention (CDC) (20). It involves an equal-weighted *quantile*-averaged combination of individual model forecasts and is not performance-weighted (19). While this quantile-averaged equal-weighted ensemble generally outperforms many of the individual models and has greater performance consistency, (21, 22) performance-weighted combinations have been demonstrated to consistently outperform equal-weighted combinations in the expert judgment literature (23, 24). Further, quantile averaging of probabilistic forecasts forms a 90% confidence interval whose width is the average of forecasters’ 90% confidence interval widths regardless of how far apart these intervals are. Thus, if one forecast has a confidence interval of 0 to 1 with a median of 0.5 while a second has a confidence interval of 9 to 10 with a median of 9.5, the quantile average yields a confidence band of 4.5 to 5.5 with a median of 5. This leads to poor statistical accuracy in over half of the cases and is not reflective of what either forecaster believes (23, 25). Other COVID-19 related studies have previously commented on the value of using the full distributions of forecasts (14, 15). These issues deserve consideration in the development of any ensemble forecasting models.

To address these research gaps, our analysis compared model forecasts of COVID-19 mortality for states within the United States with subsequent observations using several measures that reflect predictive (i.e., accuracy and precision) and probabilistic (i.e., statistical accuracy [or, as we call it, calibration] and information) performance. We also constructed three ensemble models, one equal-weighted and two performance-weighted, all of which averaged forecast probability densities, and compared their performance with each other and with the best performing individual models. Lastly, we explored whether the available models tend to provide better forecasts under certain circumstances (i.e., differing case rates, racial demographics, and forecast time periods/phases of the pandemic).

## 2. Results

### 2.1. Performance Criteria

Two aspects of performance were evaluated for each model/ensemble– (i) predictive performance (i.e., the accuracy and precision of the model/ensemble’s central estimates, or predictions); and (ii) probabilistic performance (the modeler’s ability to appropriately characterize the uncertainty in their predictions, reflected by their reported confidence intervals).

First, to evaluate the performance of a model’s predictions, each observation, O_i_, was divided by the corresponding prediction, P_m,i_, provided by the model, m, to obtain the accuracy ratio, R_m,i_ – where i indicates the question, or item, which reflects the date, state, and time range of interest:

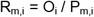

The distribution of the resulting accuracy ratios was then examined. For each model, the geometric mean (GM) and geometric standard deviation (GSD), based on the accuracy ratios for all items, N, were computed and used as respective measures of the observed accuracy and precision of the model’s predictions.

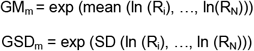

Second, to evaluate each model’s ability to appropriately characterize the uncertainty in its predictions, their probabilistic performance was assessed using the Classical Model (CM) method (10). The CM assesses ‘statistical accuracy’, or ‘calibration’, C, using Shannon’s relative information statistic, Is, which compares the forecasted probability distribution for each item with the corresponding observation (10, 26). Each observation may fall within six inter-quantile intervals, based on the five uncertainty distribution quantiles (i.e., 5%, 25%, 50%, 75%, 95%) provided by each model for every item. The calibration score for each model, C_m_, takes the following formula:

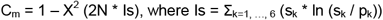

where X^2^ is the chi-square distribution function with five degrees of freedom (based on the number of quantiles), s_k_ is the percentage of observations falling in the specific inter-quantile interval, k, and p_k_ is the inherent probability of the k interval (10, 26). As the divergence between s_k_ and p_k_ increases, Is increases and C moves toward 0 (the highest calibration score possible is 1). 2N * Is, where N corresponds to the total number of items, is the log likelihood ratio which is asymptotically chi-square distributed assuming the observations are independent (10, 26). C_m_ is therefore the classical chi-square goodness of fit statistic for testing multinomial hypotheses (10, 26).

The CM assesses ‘information’, Inf, by comparing the width of the confidence intervals provided by each model with the ‘intrinsic range’ of each item (10, 26). The intrinsic range for an item is defined as the difference between the largest of the forecasted and observed values and the smallest of the forecasted and observed values (10, 26). This range is then expanded slightly by multiplying it by a user-defined expansion factor, 1+ F, where F is typically a small fraction (for our data, the standard 10% was used) (10, 26). Using this framework, the information score for each model and each item is defined as:

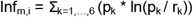

where r_k_ is the probability from a uniform (or log-uniform) probability density function over the intrinsic range (10, 26). Models which concentrate their forecasts in a narrow range will have high information scores. Therefore, while a precision score of 1 is optimal (i.e., all predictions are equally close to each other relative to the observation), the higher the information score, the better (i.e., the tighter the forecast distribution, the more information the forecast provides). See ***SI appendix*, Note 1** for more details on the CM.

To illustrate the varying model performances, **Fig. 1** provides a forest plot of forecasts reported by the individual models, as well as the constructed ensembles, during the summer 2020 period. A forest plot for the winter 2021 period is provided by ***SI appendix*, Fig. S1**.

**Fig. 1:**
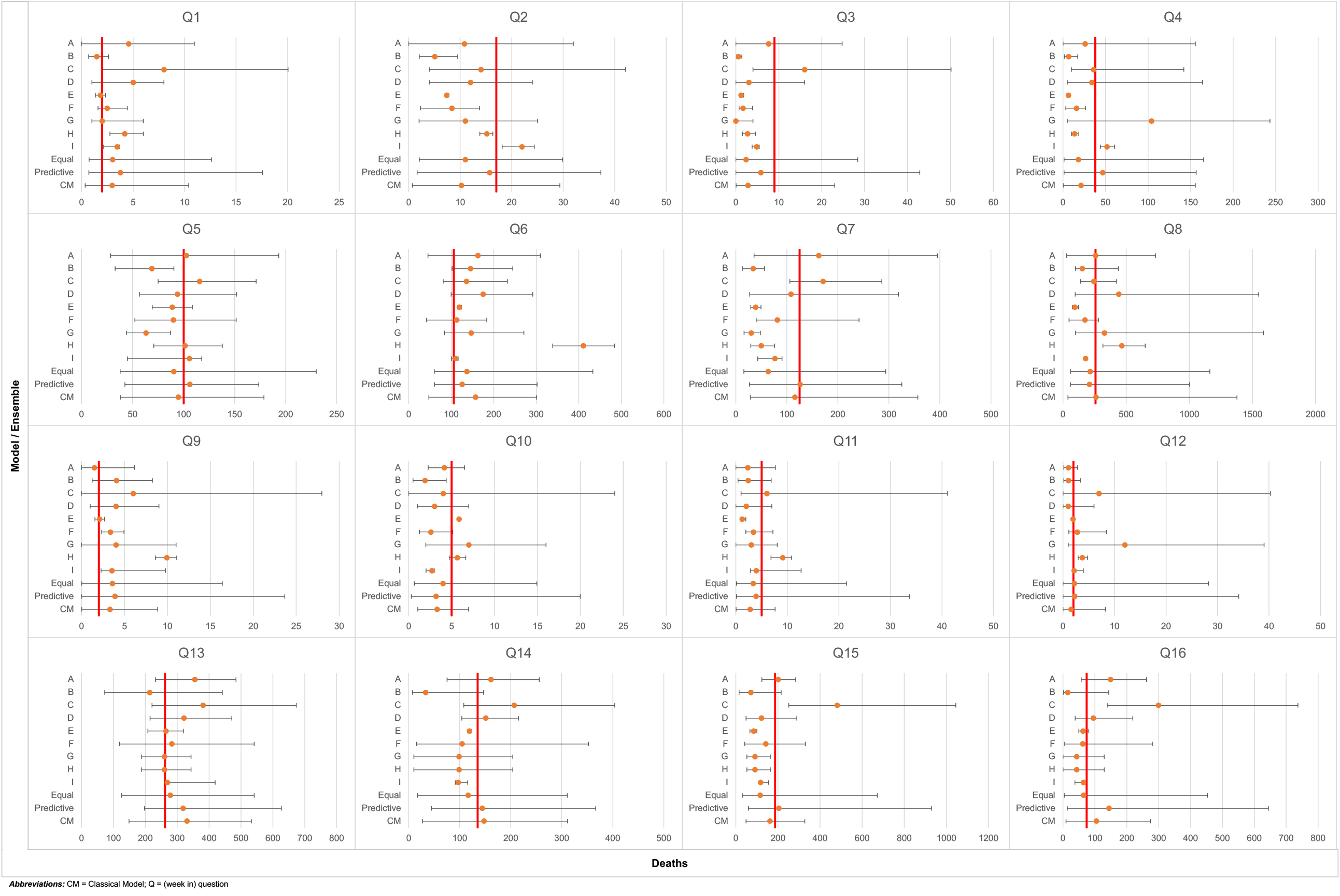
A forest plot of forecasts provided by the individual models and constructed ensembles for each of the variables of interest for summer 2020. The horizontal axis represents deaths during the week in question, the true values are given by the red vertical lines, the error bars indicate the 5^th^ and 95^th^ percentiles, and the dots represent the model’s predictions. A forest plot of forecasts for winter 2021 is provided in ***SI appendix*, Fig. S1**.

### 2.2. Individual Model Performance

The predictive and probabilistic performance results for each individual forecasting model are summarized in **Fig. 2** for both the summer 2020 and winter 2021 time periods. More quantitative detail on the performance results is provided in ***SI appendix*, Table S1**.

**Fig. 2.**
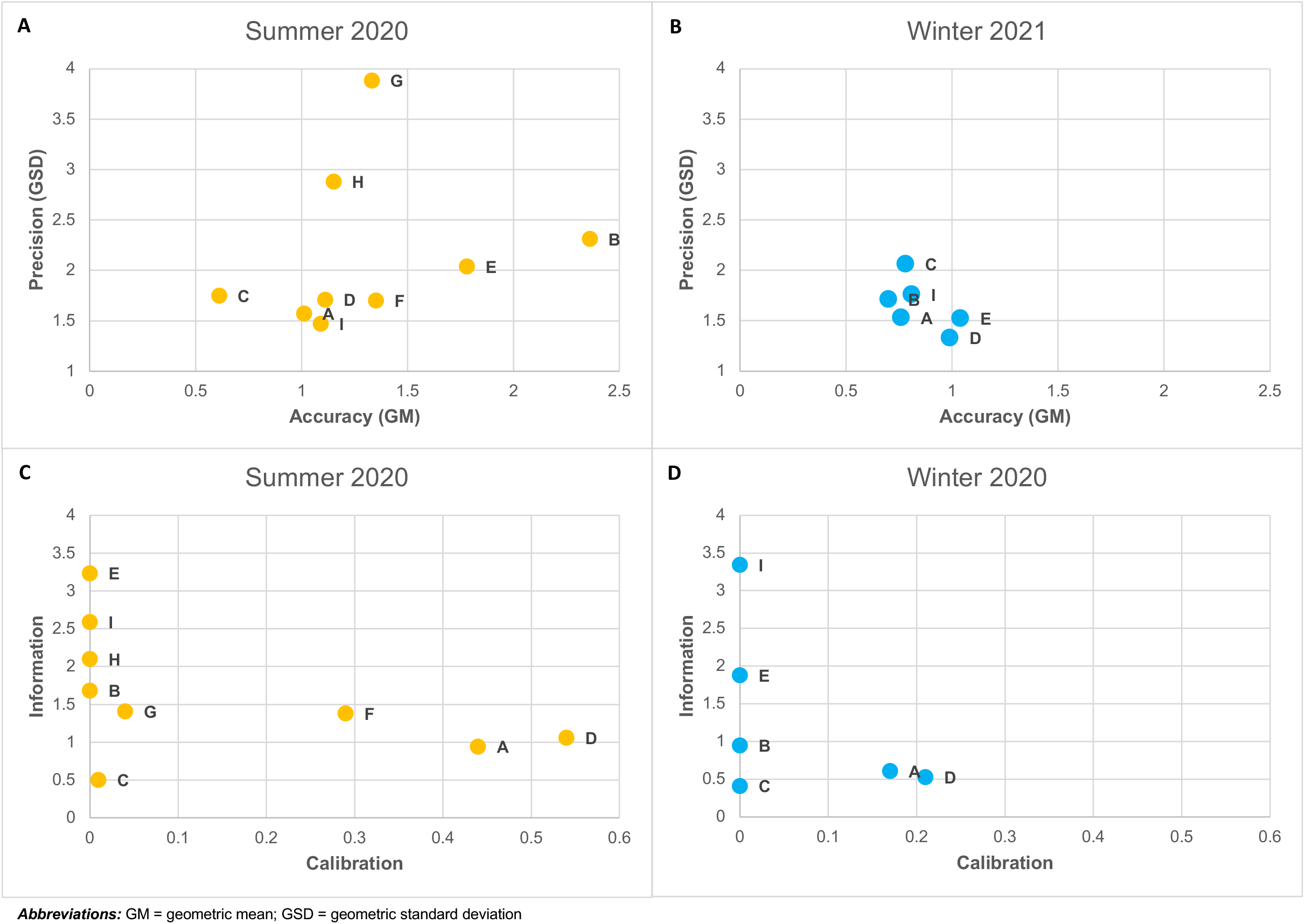
Accuracy (GM) vs. precision (GSD) of the central estimates provided by the individual models for summer 2020 (*panel A*) and winter 2021 (*panel B*) periods. Calibration vs. information scores based on the forecast densities provided by the individual models for the summer 2020 (*panel C*) and winter 2021 (*panel D*) periods. The optimal accuracy, precision, and calibration scores are 1, while the higher the information score the better.

Focusing initially on the predictive performance of the models during the summer 2020 period, we see that – (i) all models, except model C, had a GM greater than 1, indicating a systemic underprediction of COVID-19 mortality, (ii) typically model predictions had little bias when compared with the observation (≤ 35%) and precision within a factor of 2, (iii) models A, D, and I had excellent accuracy (GM = 1.01, 1.11, and 1.09, respectively) and good precision (GSD = 1.57, 1.71, and 1.47, respectively), and (iv) models B, C, and E had substantial bias (> 0.35%) and models B, E, G, and H had far worse precision (> 2). When focusing on the probabilistic performance of models during this period, only three (or perhaps, four) of the nine models considered performed at all well – models A, D, F, and to a lesser extent, G. The models that had the highest information scores (models B, E, H, and I) all had extremely low calibration scores (<< 0.01), suggesting ‘overconfidence’ – i.e., that their stated confidence intervals were far too narrow while simultaneously poorly capturing the true value.

Looking at the predictive performance during the winter 2021 period, we see that – (i) models F, G, and H no longer provide forecasts, (ii) all six remaining models, except model E, had a GM less than 1, indicating systemic overprediction of mortality, (iii) the remaining models again had little bias (< 30%) with precision within a factor of 2, except model C, (iv) model D exhibited even better accuracy (GM = 0.99) and precision (GSD = 1.33) than it did in the summer 2020 period, and model E now had excellent accuracy (GM = 1.04) and better precision (GSD = 1.52), (v) model A had good precision (GSD = 1.53) but so-so accuracy (GM = 0.76), and (vi) the models generally had better accuracy and precision than the summer 2020 period. For the probabilistic model performance, models A and D exhibited the best calibration scores (0.17 and 0.21, respectively), and again models B, E, and I were informative, but their calibration was incredibly poor (<<0.01). Interestingly, unlike accuracy and precision, model calibration and information scores generally seemed to be worse during the winter period.

The random expert hypothesis was tested to assess whether the putative differences in performance between models was due to noise. This hypothesis was rejected for both periods of assessment. More detail on this analysis can be found in ***SI appendix*, Note 2**.

Additional sensitivity analyses were conducted to evaluate whether the pervasive overconfidence in individual model forecasts was due to our selection of states with extreme case rates (i.e., only states with high or low case rates; no states with middling case rates). We carried out two sensitivity analyses, where – (i) forecasts for four states with middling case rates were added to the main dataset, and (ii) forecasts for six states with middling case rates were analyzed on their own. For these sensitivity analyses, it was expected that, if the models were sensitive to state selection by case rate, model forecasts would exhibit more appropriate confidence and substantially improve in performance. However, as illustrated by ***SI appendix*, Fig. S2-5**, while some models for summer 2020 demonstrated slight improvements in performance and exhibited slightly less confidence (i.e., lower information scores), overconfidence was still present in the same models and all models demonstrated declines in performance for the winter 2021 period. In fact, for winter 2021, all models, except Model D, had increased confidence (i.e., higher information scores) while calibration scores either remained very low (i.e., <0.01) or declined to insignificance (i.e., <0.05).

### 2.3. Ensemble Model Designs

It is possible that better and more robust estimates might be obtained by producing an ensemble based on weighted combinations of the forecasts provided by the individual models. Two approaches of potential interest were – (i) equal-weighted combinations, and (ii) performance-weighted combinations. In addition to a density-averaged equal-weighted ensemble, two density-averaged performance-weighted ensembles were considered, based on – (a) ‘predictive-performance’ weights and (b) CM weights.

For ‘predictive-performance’ weighting, each random variable from each individual model was weighted in inverse proportion to the model’s predictive variance (i.e., inverse-variance weighting the distribution of accuracy ratios of observations to predictions for each individual model):

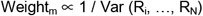

This weighting method is based on, but is not to be confused with, the inverse-variance weighting method commonly used in meta-analysis (27).

For CM weighting, based on the theory of proper scoring rules and in accordance with the CM method, performance weights were calculated as the product of both calibration and information scores (10, 26). The CM-weighted ensemble also used a threshold significance level of 0.05 (i.e., a model with a calibration score that is less than 0.05 was unweighted) for its normalized CM-weights to gain robustness without significant loss in performance (13, 26). More detail on combining “experts” (or, in this case, models) and why this cutoff was used is available in ***SI appendix*, Notes 3 and 4**.

The equal-weighted ensemble’s forecasts were established by equally weighting the probability densities of individual model forecasts, not their quantiles, to obtain aggregate forecasts. Meanwhile, for both the predictive-performance-weighted and the CM-weighted ensembles, we applied their normalized model performance weights to the probability densities of the individual model forecasts to obtain aggregate performance-based forecasts.

### 2.4. Ensemble Model Performance

The predictive and probabilistic performance results for each of the density-averaged ensemble models are summarized in **Table 1** for both the summer 2020 and winter 2021 time periods.

**Table 1.** Ensemble model performance for summer 2020 (*Top*) and winter 2021 (*Bottom*)

| Ensemble | Summer 2020 |  |  |  |  |
| --- | --- | --- | --- | --- | --- |
|  | Accuracy<br>- GM | Precision<br>- GSD | Calibration | Information | CM Weight<br>(Unnormalized) |
| Equal Weight | 1.24 | 1.60 | 0.03 | 0.72 | 0.02 |
| Performance –<br>Predictive<br>Weight | 0.91 | 1.41 | 0.04 | 0.55 | 0.02 |
| Performance –<br>CM Weight* | 1.12 | 1.57 | 0.54 | 0.81 | 0.43 |

| Ensemble | Winter 2021 |  |  |  |  |
| --- | --- | --- | --- | --- | --- |
|  | Accuracy<br>- GM | Precision<br>- GSD | Calibration | Information | CM Weight<br>(Unnormalized) |
| Equal Weight | 0.84 | 1.55 | 0.17 | 0.36 | 0.06 |
| Performance –<br>Predictive<br>Weight | 0.84 | 1.49 | 0.25 | 0.35 | 0.09 |
| Performance -<br>CM Weight* | 0.85 | 1.45 | 0.44 | 0.43 | 0.19 |

\* A hypothesis rejection significance level of 0.05 was used for this ensemble. More detail on why this was used is available in **SI appendix, Notes 3 and 4.**
**Abbreviations:** CM = Classical Model; GM = geometric mean; GSD = geometric standard deviation

Both performance-weighted ensembles would be expected to outperform the equal-weighted ensemble (23, 24) and they did fulfill this expectation on almost all measures of performance, however, the differences in predictive performance were often small. The accuracy of the performance-based combinations was appreciably better than the equal-weighted combination in the summer 2020 data, but this advantage was not seen in the winter 2021 data. Very small differences in precision were observed in both data sets.

To explore whether these differences in accuracy between the summer 2020 and winter 2021 periods were because three of the nine models included in the summer 2020 analysis were not reflected in the winter 2021 data, we reassessed model performance in the summer of 2020 using only data from the six models that were available during both seasons. The differences in accuracy seen in the summer of 2020 between the equal-weighted and performance-weighted combinations were even greater (equal weight = 1.23, predictive weight = 0.94, CM unnormalized weight = 1.02) than those observed in the full data set. More detail on this analysis is provided in ***SI appendix*, Table S2**.

Though predictive performance differences were small, there were substantial differences in probabilistic performance. The calibration scores of the CM-weighted ensemble (0.54 in summer 2020; 0.44 in winter 2021) were far better than those of the equal-weighted ensemble (0.03 during the summer, 0.17 during the winter) or the predictive-performance-weighted ensemble (0.04 during the summer, 0.25 during the winter). There were also small differences in information scores, where again the CM-weighted ensemble slightly outperformed the other two ensembles – resulting in substantially higher CM weights during both periods.

### 2.5. Performance by Domain

We also compared the performance of the individual models across three domains of interest – (i) race (i.e., states which are heavily non-Hispanic White vs. states with relatively large non-Hispanic Black populations), (ii) COVID-19 case rates (i.e., states with a relatively low amount of weekly cases per 100,000 population vs. states with a relatively high amount of weekly cases per 100,000 population), and (iii) forecast period (i.e., forecasts of mortality for the upcoming week vs. forecasts of mortality for the week ending four weeks from the date on which the forecast was made). The performance of the equal-weighted density-averaged ensemble stratified by these three different domains (i.e., eight forecasts per domain) is provided in **Table 2**.

**Table 2:** Equal-weighted ensemble model performance for summer 2020 (*Top*) and winter 2021 (*Bottom*) stratified by different domains.

| Sub-Domain | Summer 2020 |  |  |  |  |
| --- | --- | --- | --- | --- | --- |
|  | Accuracy<br>- GM | Precision<br>- GSD | Calibration | Information | CM Weight<br>(Unnormalized) |
| High % non-Hispanic White<br>(ID, ME; VT, WY) | 1.30 | 1.86 | 0.60 | 0.86 | 0.52 |
| High % non-Hispanic Black<br>(LA, NY; GA, MD) | 1.19 | 1.34 | 0.05 | 0.59 | 0.03 |
| High Case Rate<br>(ID, LA; VT, GA) | 1.42 | 1.77 | 0.32 | 0.67 | 0.21 |
| Low Case Rate<br>(ME, NY; WY, MD) | 1.09 | 1.39 | 0.14 | 0.78 | 0.11 |
| One Week Ahead | 0.95 | 1.41 | 0.27 | 0.53 | 0.14 |
| Four Weeks Ahead | 1.62 | 1.56 | 0.05 | 0.92 | 0.05 |

| Sub-Domain | Winter 2021 |  |  |  |  |
| --- | --- | --- | --- | --- | --- |
|  | Accuracy<br>- GM | Precision<br>- GSD | Calibration | Information | CM Weight<br>(Unnormalized) |
| High % non-Hispanic White<br>(ID, ME; VT, WY) | 0.69 | 1.56 | 0.14 | 0.36 | 0.05 |
| High % non-Hispanic Black<br>(LA, NY; GA, MD) | 1.02 | 1.42 | 0.73 | 0.37 | 0.27 |
| High Case Rate<br>(ID, LA; VT, GA) | 0.81 | 1.76 | 0.66 | 0.41 | 0.27 |
| Low Case Rate<br>(ME, NY; WY, MD) | 0.87 | 1.34 | 0.16 | 0.31 | 0.05 |
| One Week Ahead | 0.88 | 1.51 | 0.02 | 0.35 | 0.01 |
| Four Weeks Ahead | 0.80 | 1.62 | 0.06 | 0.37 | 0.02 |

**Abbreviations:** CM = Classical Model; GM = geometric mean; GSD = geometric standard deviation; GA = Georgia; ID = Idaho; LA = Louisiana; MD = Maryland; ME = Maine; NY = New York; VT = Vermont; WY = Wyoming **Note:** Analogues of **Table 2** for the CM- and predictive-performance-weighted ensembles are also provided in **SI appendix, Table S7 and S8**, respectively.
**Abbreviations:** GM = geometric mean; GSD = geometric standard deviation

Both the accuracy and precision of predictions for states with relatively high non-Hispanic Black populations were better than for states with a largely non-Hispanic White population. This difference appeared to be stable and was seen in both the summer 2020 and winter 2021 data. However, there is no evidence of any stable differences in the calibration and information scores of forecasts depending on the racial composition of the state.

The summer 2020 data suggests that predictions were more accurate and precise in states with low case rates, and this remained true in the more recent winter 2021 data, but the difference in accuracy is not as substantial. On the other hand, model calibration scores were somewhat better in states with high case rates. This difference appeared to be stable and was seen with both the summer 2020 and winter 2021 data.

Lastly, as expected, the predictions of deaths in the near-term appear to be more accurate and precise than those in the mid-term, especially for the summer 2020 data. This remained true for the more recent winter 2020 data, but the differences in both accuracy and precision were smaller. There is some evidence that calibration scores were better for near-term forecasts in the summer data, but this did not persist in the winter data.

## 3. Discussion

There has been little published peer-reviewed information regarding the performance of COVID-19 forecasting models, and what is available has focused mostly on the performance of their central estimates (i.e., predictive performance). Very little has been reported on the performance of their uncertainty estimates (i.e., probabilistic performance), which could inform users how often they would be surprised by observations outside of a model’s forecasted confidence intervals.

To address this gap in knowledge, we borrowed from the formally elicited expert judgment literature to demonstrate a commonly used approach for resolving this issue (i.e., the Classical Model; CM). We then evaluated both the predictive (i.e., accuracy and precision) and probabilistic (i.e., calibration and information) performance of COVID-19 forecasting models during two independent eight-week time periods (i.e., summer 2020 and winter 2021). In our analysis, some models exhibited good predictive performance but were found to have poor probabilistic performance (e.g., model I, and for the winter, E), and vice versa. Of the nine selected forecasting models, only two models, A and D, did reasonably well in both predictive and probabilistic performance, with model D demonstrating the most consistent dominance in all performance measures.

This study was the first to utilize the CM method with COVID-19 forecasting models to measure their probabilistic performance and these CM metrics provided valuable insights. The fact that all but three of the nine models considered (models A, D, and F) had such low calibration scores for both time periods and that most of those also had high information scores indicates pervasive overconfidence.

There is strong evidence that most expert judgments are overconfident, (15, 28, 29) that is why there are often elaborate procedures to minimize this in expert elicitations (30). It is understandably difficult to predict changes in human behavior, and, while rare, there are occasional anomalies in reported data (e.g., large revisions or bulk reporting) that may further bias a model’s forecast. This issue has been highlighted in other assessments of these models (22, 31). While, to the best of our knowledge, there were no such anomalies demonstrated in the observed data utilized for this study, it is clear from these results and the literature that many modelers need to carefully re-evaluate how they are quantifying uncertainty in their predictions.

We found that the tendency of models to underpredict in the summer of 2020 was replaced by a tendency to overpredict in the winter of 2021 – suggesting that model predictions lag the actual changes in disease rates, which were generally increasing in the summer of 2020 and generally decreasing in the winter of 2021. This relative unresponsiveness to more rapid shifts has been well documented by the COVID-19 Forecast Hub and their studies (7, 22). This also could be the reason why model forecasts generally performed worse during winter 2021 versus summer 2020, as many states experienced rapid and substantial declines in weekly mortality counts during late winter 2021.

All the assessed individual models were variations of a susceptible-exposed-infectious-removed (SEIR) compartmental model (32-40). They all forecasted at the U.S. state level, and some also forecasted at the national and county levels (8). Every model provided forecasts at the daily scale, which could then be aggregated to the weekly scale for at least four weeks ahead from forecast date (8).

However, while all models used mortality data to inform compartment transition rates, they varied in how they incorporated case, hospitalization, demographic, and mobility data (32-40). Both models A and D utilized case and mortality data, and model A utilized demographic data, but neither model incorporated hospitalization nor mobility data (32, 35). Using case and mortality data, models A and D extrapolated deaths from lagged case counts, with a 14-day lag for model A (32) and a varying 15-to 30-day lag for model D (35). Additionally, while models A and D, like most other models, incorporated time-varying COVID-19 transmission rates which could indirectly reflect social distancing measures, neither model directly incorporated changes in social distancing measures for forecasts in the near-term (although, model A may have incorporated changes in social distancing measures for forecasts past three weeks) (32, 35).

The designs of these forecasting models have also not been static. Modeling groups have been learning and adapting their models over the course of the pandemic. Thus, a model’s forecasts for the summer and winter are broadly comparable, but its methods to produce the forecasts are not necessarily identical. This further emphasizes the significance of our integration of expert judgment methods, as we essentially assessed the performance of ‘judgments’ from model forecasting groups rather than from the models themselves, which are ever evolving.

For both time periods, we constructed and evaluated a density-averaged predictive-performance and CM-performance-weighted ensemble, which were respectively based on the predictive and probabilistic performance of the individual forecasting models. Both the performance-weighted ensembles outperformed the equal-weighted ensemble, although the differences in predictive performance were modest. This result is anticipated in the Cooke, Marti, & Mazzuchi (2021) study (16). In contrast, when evaluated in terms of probabilistic performance there were clear advantages to performance weighting using the CM weights. While a density-averaged equal-weighted ensemble may still hold merit (e.g., improved performance scores over many individual models; does not require prior forecasts/realizations to train the ensemble; easy to produce), for best results, particularly when probabilistic performance is considered, CM performance-weighting should be utilized. Additionally, the CM method removes from aggregation those models that are not able to appropriately characterize the uncertainty in their predictions, and thus should not be relied upon. In any case, if performance-based modeling is not desired, ensembles should at least be constructed based on averaged densities and not quantiles bins.

Across the three domains and two performance attributes considered, consistent and substantial performance differences were seen only in a few instances – and these differed depending on whether one focuses on predictive or probabilistic performance. Predictive performance was generally better for forecasts in states with relatively high non-Hispanic Black populations and relatively low case rates, but the observed differences were often modest. Probabilistic performance was consistently better when case rates were high. This difference in performance based on case rates could be explained by models having difficulty adapting to rapid shifts (reflected in states with high case rates), but this added uncertainty may be more appropriately reflected in their forecasts. Finally, as expected based on previous study findings (9, 31) and the disease progression timeline (i.e., cases now strongly predict deaths in 17-21 days), (41) near-term predictions (i.e., deaths in the next week) outperformed mid-term predictions (i.e., deaths in the week ending four weeks from forecast date). However, we would like to emphasize that these results are not definitive and may not be representative of the results that would be based on all models and all states.

One study, currently in preprint, also evaluated the predictive and probabilistic accuracy of individual models (using different metrics) and found that accuracy degraded as models made predictions further into the future and that individual models were frequently overconfident (31). The authors also found that the models with few data inputs were among the most accurate and the same models demonstrated superior performance in their analysis (31). Thus, many of their findings agreed with ours.

It is also important to note that the CM is not the only protocol that analyzes and aggregates expert judgments (or, in this case, model forecasts). Although other expert judgment protocols that typically involve behavioral aggregation could not be utilized in this post hoc analysis, (17, 18) it would be interesting to elicit from modeling groups a “best” model design and this could be the focus of a separate paper.

Our sense is that these results are more suggestive than conclusive, because – (i) due to the selection criteria, only nine models were examined, and three of these ceased forecasting by the winter 2021 period; (ii) they are based on model performance in one eight-week period in the summer of 2020 and one eight-week period in the winter of 2021; (iii) they are limited to eight states (although more states were examined for sensitivity in ***SI appendix*, Fig. S2-S5**); and (iv) our analysis did not attempt to explain what model design choices led to better or worse performance.

If we were to reexamine the data, we would have analyzed all models that met our criteria for the winter period, not just those that were analyzed for the summer and continued forecasting into the winter. Additionally, in the expert judgment literature, 8 to 15 calibration variables are standard and any more is thought to be unnecessary (23). Thus, we believe this analysis to be an adequate demonstration of the CM for the selected models for those selected time periods and states. However, we understand that, considering the limited scope of this analysis, and with the situation and the modeling groups’ ability to forecast always changing, for more definitive conclusions on the performance of the specific individual models, a broader evidence base should be utilized. Excalibur, the program that was used to run the CM, was designed for the analysis of formally elicited expert judgment with limited calibration questions (42). In the future, we hope to adapt the Excalibur code so that it can handle the full set of data available for the evaluation of COVID-19 models.

Conversely, our analysis has several strengths – (i) it evaluated the performance of a set of leading models which have been used to forecast COVID-19 mortality in the US; (ii) it considered *both* predictive (i.e., accuracy and precision) and probabilistic (i.e., calibration and information) performance, and was the first study to utilize the CM method to accomplish this; (iii) it assessed several approaches for constructing *density-averaged* ensemble models, and the CM-weighted ensemble exhibited superior performance; (iv) it examined performance differences across states selected to reflect differences in racial composition and variations in COVID-19 case rate; and (v) it considered data from two distinct phases of the pandemic.

We conducted this analysis to demonstrate the limitations of performance evaluations which consider only predictive performance. While we believe this analysis of individual COVID-19 forecasting models to be adequate, considering its limited scope and the dynamic model designs, we do not believe that our results provide definitive conclusions about the performance of the specific models considered. Rather, we suggest the availability, utility, and importance of an approach that has been used for years in the expert judgment literature for characterizing the probabilistic performance of such forecasts and constructing high performing ensembles. We believe this application demonstrates that the CM could be a useful methodology not just for COVID-19 modeling but other applications as well. We hope that users will find this approach to be valuable in their efforts to better characterize and communicate forecast performance.

## 4. Materials and Methods

Our analysis involved a comparison of model forecasts with subsequent observations of weekly deaths from COVID-19 in four states (Idaho, Louisiana, New York, and Maine) over an eight-week period during the summer of 2020 and four other states (Georgia, Maryland, Vermont, and Wyoming) over a subsequent eight-week period during the winter of 2021.

The states considered in our analysis were selected based on recent case rates of COVID-19 (cases/100,000 population within the previous week) (43) and racial composition (majority non-Hispanic Black vs. majority non-Hispanic White) (44). Changes in COVID-19 case rates were of interest as they precede changes in COVID-19 death rates by a period of about 18.5 days (41). Racial composition was of interest as the COVID-19 mortality rate for non-Hispanic Black Americans at the time of analysis was more than twice that of non-Hispanic White Americans, and this disparity persisted even when accounting for other demographic and socioeconomic factors (45).

With these two domains in mind, our goal was to assess two states for each time period with relatively high case rates (Idaho and Louisiana; Georgia and Vermont); two with relatively low case rates (Maine and New York; Maryland and Wyoming); two with a relatively high fraction of population reported as non-Hispanic Black (Louisiana and New York; Georgia and Maryland); and two with a relatively high fraction of population reported as non-Hispanic White (Idaho and Maine; Vermont and Wyoming). This was done to assess how models may perform forecasting for states under varying circumstances. More detail on how the case rates and racial composition for states were determined, as well as how states were selected, is available in ***SI appendix*, Note 5** and **Tables S3 and S4**.

We were also interested in the models’ ability to forecast COVID-19 deaths in both the near-term and the medium-term. Near-term performance was gauged using projected COVID-19 deaths in the week immediately after the forecast was made. Medium-term performance was gauged using projected COVID-19 deaths in the week ending four weeks after the forecast was made.

Our evaluations of model performance for the eight states and the two forecast periods of interest (week ending one week in the future and week ending four weeks in the future) were examined twice within each period – once for forecasts made on June 13^th^, 2020, or January 10^th^, 2021, and a second time for forecasts made on July 11^th^, 2020, or February 7^th^, 2021 (no overlap in forecasts). In total, 16 comparisons of model forecasts with observed deaths were made for each time period and each model.

Of the many models providing data to the COVID-19 Forecast Hub’s data repository (8), only the models that provided weekly COVID-19 mortality forecasts made on the same day of the week, with forecasts at the weekly timescale for all four of the initially assessed states and without missing forecasts were selected for this analysis. These include: OliverWyman-Navigator (Model A) (32), MOBS-GLEAM_COVID (Model B) (33), JHU_IDD-CovidSP (Model C) (34), UMass-MechBayes (Model D) (35), UCLA-SuEIR (Model E) (36), YYG-ParamSearch (Model F) (37), UT-Mobility (Model G) (38), USACE-ERDC_SEIR (Model H) (39), and Covid19Sim-Simulator (Model I) (40). To understand whether model performance was unique to one phase of the pandemic or whether performance was consistent across the two phases, these same models were assessed during the winter 2021 period.

## Data Availability

Observed state COVID-19 mortality and case data was gathered from the Centers for Disease Control and Prevention (CDC) (43). State population and racial composition data was collected from one-year estimates from the Census Bureau's 2018 and 2019 American Community Survey (ACS) (44). For racial composition statistics and case rate data, please see SI appendix, Tables S3 and S4.
Model forecasting data was gathered from the COVID-19 Forecast Hub's publicly available structured data storage repository on GitHub (8). For the model and ensemble predictions, their uncertainty distributions, and the subsequent observations of COVID-19 mortality, please see SI appendix, Tables S5 and S6.

## Code Availability

Data was analyzed using Microsoft Excel and EXCALIBUR (a software package for using the Classical Model) (42).

## Data Availability

Observed state COVID-19 mortality and case data was gathered from the Centers for Disease Control and Prevention (CDC) (43). State population and racial composition data was collected from one-year estimates from the Census Bureau’s 2018 and 2019 American Community Survey (ACS) (44). For racial composition statistics and case rate data, please see ***SI appendix*, Tables S3 and S4**.

Model forecasting data was gathered from the COVID-19 Forecast Hub’s publicly available structured data storage repository on GitHub (8). For the model and ensemble predictions, their uncertainty distributions, and the subsequent observations of COVID-19 mortality, please see ***SI appendix*, Tables S5 and S6**.

## Acknowledgments

We want to thank Willy Aspinall, Jouni Tuomisto, and Jacqueline Macdonald for contributing to our thoughts about this and for their feedback on our early drafts of the paper.

