## Supplementary material for "Post Hoc Evaluation of Probabilistic Model Forecasts: A COVID-19 Case Study": SI Appendix

**This PDF file includes:**

Supplementary Text (Notes 1 through 6)

Figures S1 to S5

Tables S1 to S12

SI References

**Supplementary Information Text**

**Contents**

1. **Technical Details of the Classical Model**
2. **Random Expert Hypothesis**
3. **Combining Experts**
4. **Robustness Analysis**
5. **Determining State Racial Composition and COVID-19 Case Rates**
6. **Supplemental Figures & Tables**

1. **Technical Details of the Classical Model**

The current study considered five assessed quantiles (i.e., 5%, 25%, 50%, 75% and 95%) for each elicited uncertain quantity, or item. The expert could be a human assessor, a computer code, or some combination of the two.

1.1. **Statistical Accuracy (aka Calibration)**

The assessed q­­uantiles divide the range of possible values into six inter-quantile intervals for which an expert’s probabilities are known (e.g., p_1_ = 0.05 [less than or equal to the 5% quantile]; p_2_ = 0.20 [greater than the 5% quantile and less than or equal to the 25% quantile], etc.). If N quantities are assessed as calibration questions, or items, each expert may be regarded as a statistical hypothesis, namely that each realization falls in one of the six inter-quantile intervals with the probability vector:

p = (0.05, 0.20, 0.25, 0.25, 0.20, 0.05)

Suppose we have realizations x_1_, …, x_N_ of these quantities. We may then form the sample distribution of the expert's inter-quantile intervals as s_e_ = (s_1,e_, s_2,e_, ..., s_6,e_), where:

s_1,e_ = # {i | x_i_ ≤ 5% quantile} / N

s_2,e_ = # {i | 5% quantile < x_i_ ≤ 20% quantile} / N

...

s_6,e_ = # {i | 95% quantile < x_i_} / N

Note that the sample distribution depends on the assessments from the expert, e. If the realizations are indeed drawn independently from a distribution with quantiles as stated by the expert, then the quantity:

$2N\sum_{i=1}^{6} s_{i}\ln\left( \frac{s_{i}}{p_{i}} \right)$

(1.1)

is asymptotically distributed as a chi-square random variable with five degrees of freedom. This is the so-called likelihood ratio statistic. If we extract the leading term of the logarithm, we obtain the familiar chi-square test statistic for goodness of fit. If, after a few realizations, the expert was to see that all realizations fell outside of their 90% central confidence interval, they might conclude that their intervals were too narrow and broaden them on subsequent assessments. This means that, for this expert, the uncertainty distributions are *not* independent, and they learn from the realizations. Expert learning is not a goal of a study, and their joint distribution is not elicited. Rather, the analyst wants experts who do not need to learn from the elicitation. Hence the analyst scores the expert, e, as the statistical likelihood of the hypothesis:

*H_e_: The inter-quantile interval containing the true value for each item is drawn independently from probability vector, p.*

A simple test for this hypothesis uses the test statistic (1.1), and the likelihood, or p-value, or **statistical accuracy score** (aka **‘calibration score’**) of this hypothesis, is:

C_e_ = P (χ ≥ r | H_e_)

(1.2)

where χ is a chi-square distributed random variable with five degrees of freedom and r is the value of (1.1) based on the observed values x_1_, …, x_N_. It is the probability under hypothesis H_e_ that a deviation at least as great as r should be observed on N realizations if H_e_ were true. Calibration scores are absolute and can be compared across studies. However, before doing so, it is appropriate to equalize the power of the different hypothesis tests by equalizing the effective number of realizations. To compare scores on two data sets with N and N’ realizations, we simply use the minimum of N and N' in (1.1), without changing the sample distribution, s.

Although the calibration score employs the language of simple hypothesis testing, it must be emphasized that we are not rejecting expert-hypotheses; rather we are using this language to measure the degree to which the data supports the hypothesis that the expert's probabilities are statistically accurate. Low scores, near zero, mean that it is unlikely that the expert’s probabilities are correct.

1.2. **Information**

Measuring information requires associating a density to each assessment for each item. To do this, we use the unique density that complies with the expert's quantiles and is minimally informative with respect to a background measure. For a uniform background measure, the density is constant between the assessed quantiles, and is such that the total mass between the quantiles agrees with p. The background measure is not elicited from experts as it must be the same for all experts; instead, it is chosen by the analyst. The Classical Model (CM) uses by default either the uniform or the log-uniform background measure.

The uniform and log-uniform background measures require an ‘intrinsic range’ on which these measures are concentrated. The CM implements the so-called ‘k% overshoot rule’ (i.e., for each item, we consider the smallest interval, I = [L, U], containing all the assessed quantiles of all experts and the realization, if known). This interval is extended to:

I^*^ = [L^*^, U^*^]; L^*^ = L – k (U - L) / 100; U^*^ = U + k (U - L) / 100

(1.3)

The value of k is chosen by the analyst. A large value of k tends to make all experts look quite informative and tends to suppress the relative differences in information scores. The **information score** of the expert, e, on assessments for uncertain quantities 1, …, N, is:

Inf_e_ = (1 / N) ∑_i = 1, …, N_	f_e,i_ ln (f_e,i_ | g_i_)

(1.4)

where g_i_ is the background density for item, i, and f_e,i_ is the elicited density from the expert, e, for the i. This is proportional to the relative information of the expert's joint distribution given the background, under the assumption that the items are independent. As with calibration, the assumption of independence reflects a desideratum of the analyst and is not an elicited feature of the expert's joint distribution. The information score does not depend on the realizations. An expert can earn a high information score by choosing their quantiles very close together.

Evidently, the information score of an expert depends on the intrinsic range and on the assessments of the other experts. Hence, information scores cannot be compared across studies.

The information score is a “slow” function, that is, large changes in the assessments produce only modest changes in the information score. This contrasts with the likelihood function in the calibration score (1.1)*,* which is a very fast function. This causes the normalized product of calibration and information to be driven by the calibration score. It also means that modest changes in informativeness correspond to sizeable changes in the distributions. Increasing informativeness by a factor of two roughly corresponds to halving the distance between the 95^th^ and 5^th^ percentiles.

**1.3. Brief Digression on Scoring Rules for Individual Items and Average Probabilities**

Scoring rules were introduced by de Finetti in 1937 as tools for eliciting subjective probabilities (1) and have been further developed by many authors (2). An expert receives a score as a function of their probability assessment and the realization. The score is strictly proper if the expert maximizes their expected score per item by, and only by, stating their true belief. Using a result of Murphy (1977), (3) DeGroot and Fienberg (1983) gave an additive decomposition of strictly proper rules into ‘calibration’ and ‘refinement’ terms, (4) thereby replacing Murphy’s ‘resolution’ (refinement applies only to well-calibrated experts). In the case of the logarithmic rule, refinement becomes Kullback-Leibler Divergence (aka, directed divergence, cross entropy, or relative information [we use the latter term]).

Scoring rules were not designed for combining experts and are not suitable in that role. Consider 100-coin tosses. An expert assesses the probability of heads on each toss as ½. Their score for the outcome heads is the same as their score for tails on each toss. If their score for all 100 assessments is a function of their 100 scores for the individual tosses, then their score for 100 tosses is independent of the outcome sequence (i.e., the outcome of 100 heads receives the same score as 50 heads and 50 tails). There are many such examples of this (5, 6).

To avoid problems with scores for individual items, Cooke introduced scoring rules for average probabilities (5). Let random variables X_1_, ..., X_n_ take outcomes in a finite set, O, let M_O_ be the set of probability measures on O, and let M_n_ be the set of probability measures on X_1_, ..., X_n_. For Π ∈ M_n_, let π be the vector of average probabilities, that is, π_i_ = (1 / n) Σ_j=1, ..., n_ Π (X_j_ = i). Let s be the observed relative frequency of outcomes for realization, (X_1_, ..., X_n_) = (x_1_, ..., x_n_). A scoring rule for average probabilities assigns a number, R, to the pair (π, s). R is strictly proper (positively sensed) if:

for all Π ∈ M_n_, argmax _ϕ ∈Mo_ E_Π_ (R (ϕ, s) = π.

This says, whatever the expert’s belief, Π, about (X_1_, ..., X_n_), they achieve their maximal expected score by stating the probability, π, over outcomes which corresponds to their average probabilities. The proofs are a bit more complicated because, “for all Π,” goes over a much larger set than the argmax over M_o_.

There is a representation theorem in Cooke (1991) for such rules (5). However, more useful in practice are rules which are asymptotically strictly proper as n → ∞. These rules allow the product form in the CM.

1.4. **Brief Digression on the User-Defined Expansion Factor**

Familiarity with foundations teaches that the problem of combining experts’ judgments is not a mathematical problem. The laws of probability even supplemented with Savage’s axioms and the theory of proper scoring rules, will never tell us how to combine experts. The problem is more akin to finding an optimal design in engineering. For example, a bicycle obeys Newton’s laws but doesn’t follow from them. Any working design will involve features motivated by practicalities rather than laws. So let it be with measures of “spread”.

Traditional measures like the standard deviation and prediction intervals are unsuitable because they inherit the physical dimension of the underlying variables: changing from meters to kilometers changes the numbers. To compare spreads across variables with different physical dimensions we need a measure which is scale invariant. We also need it to be “tail insensitive” because the tails in expert judgment are poorly constrained. Finally, it must be “slow”.

The theory of asymptotic proper scoring rules for average probabilities gives a product form of: measure of statistical accuracy × measure of lack of spread. Weights will be formed by normalizing such products. We want statistical accuracy (aka, calibration; a very fast function) to dominate, therefore the measure of “lack of spread” must be slow. Relative information fits the bill and requires specifying “relative to what”. This is as it should be because information in a distribution is always relative to another (background) distribution.^[1]^

Since the background distribution must integrate to one, it must be “concentrated” somewhere. The uniform or log-uniform distributions are chosen as default backgrounds because they have no location parameter other than the endpoints of their support. However, we do need to specify the compact support of the background for each item. We do this by choosing the smallest interval containing all assessments and the realization, if available, plus a k% overshoot. If k is very large then all experts appear very informative relative to the background, and this tends to discount the role of information in determining weights. The CM dictates that the choice of uniform/log-uniform and k are choices of the analyst, and any such choices must be controlled by user parameters in the code. Thus, we can see that k must be very large to make much difference. Uniform vs log-uniform can have an influence; the guidance is that, if we reason about orders of magnitude − use log-uniform.

2. **Random Expert Hypothesis**

The random expert hypothesis states that putative differences in performance between experts are just noise and do not indicate persistent differences of the experts (9). One way to test this hypothesis is to compare panel wide performance metrics in the original panel with the same metrics as generated by a large set of “scrambled panels” in which the assessments are randomly re-allocated to experts, thus wiping out any ‘expert effect’. The code used for this exercise is based on the 5%, 50% and 95% quantiles, whereas this study also used the 25% and 75% quantiles. This causes the scores to differ somewhat. Considering statistical accuracy (aka, calibration) and information, we are interested in the panel maximum, minimum and averages. **Table S9** shows, for example, that the average calibration score in the original panel was 0.01. In 62% of the 1000 scrambled panels the average calibration score was lower than 0.01. In all the scrambled panels the minimum calibration score was greater than the original panel minimum calibration. Although the scrambling was able to exceed the panel average calibration in 38% of the cases, it was never able to get scores as low as the original panel. The average information score is always the same in the scrambled and original panels, but the scrambling was unable to reproduce the highest and lowest scores. The same pattern is observed in the second panel. Hence, the random expert hypothesis cannot be supported for either panel.

3. **Combining Experts**

The ‘combined score,’ Cs, of the expert, e, is the dimensionless quantity*:*

Cs_e_ = C_e_ * Inf_e_

(2.1)

A scoring rule is (asymptotically) strictly proper if an expert achieves their (long run) maximal expected score by, and only by, giving assessments corresponding to their true beliefs. That is, an expert maximizes their long run expected score by, and only by, ensuring that the probabilities, p = (0.05, 0.20, 0.25, 0.25 0.20, 0.05), correspond to their true beliefs. The theory of proper scoring rules tells us that (2.1) becomes a proper scoring rule if it is augmented with a cutoff, α, on the calibration score such that an expert is unweighted if C < α. The α is like the significance level in simple hypothesis testing, but its purpose is different. The goal is to measure “goodness” with a strictly proper scoring rule.

A combination of expert assessments is called a ‘decision-maker’ (DM)*.* All DMs discussed here are examples of linear pooling. The Classical Model is essentially a method for deriving weights in a linear pool. A "good probabilistic expert" corresponds to an expert with good calibration (i.e., high statistical likelihood; high p-value) and high information. We want weights which reward good experts, and which pass these virtues on to the DM.

The reward aspect of weights is very important. We could simply solve the following optimization problem: find a set of weights such that the linear pool under these weights maximizes the combined score of the DM*.* When solving this problem with real data, one finds that the weights do not generally reflect the performance of the individual experts. As we do not want an expert's influence on the DM to appear haphazard, and we do not want to encourage experts to game the system by tilting their assessments to achieve a desired outcome; we must impose a strict scoring rule constraint on the weighting scheme.

The scoring rule constraint requires that the combined score is multiplied by the indicator function, 1_α_ (C_e_ ≥ α), which takes the value 1 if C_e_ ≥ α and 0 otherwise*:*

w_α,e_ = C_e_ * Inf_e_ * 1_α_ (C_e_ ≥ α)

(2.2)

This says that the expert, e, is weighted only if their calibration is at least α*.* The resulting DM is a function of α:

DM_α,i_ = ∑_e=1, ..., E_ w_α,e_ f_e,i_ / ∑_e=1, ..., E_ w_α,e_

(2.3)

Scoring rule theory does not say what the value of α should be. In practice there are three ways for choosing α*.* The ‘optimized’ DM chooses α such that the resulting combined score of the DM is maximized. The optimized DM is DM_α*_ where α* maximizes C(DM_α_) × Inf(DM_α_). This typically leads to choosing an α so high that only one or two experts are weighted. The ‘statistical threshold’ DM chooses an α to distribute weight over experts with “acceptable” calibration (typically, α = 0.05 or 0.01). The “inclusive” DM chooses an α so low that all experts are weighted. If this choice is made a posteriori, then this is not a strictly proper scoring rule. It is to be noted that unweighted experts still have influence over the intrinsic range, and their rationales are recorded.

These weights are termed global because the information score is based on all the assessed calibration items. A variation on this scheme allows a different set of weights to be used for each item. This is accomplished by using information scores for each item rather than the average information score:

w_α,e,i_ = 1_α_ (C_e_ ≥ α) * C_e_ * f_e,i_ ln (f_e,i_ | g_i_)

(2.4)

For each α we define the ‘item weight’ DM_α_ for the item, i, as:

IDM_α,i_ = ∑_e=1, ..., E_ w_α,e,i_ f_e,i_ / ∑_e=1, ..., E_ w_α,e,i_

(2.5)

The same variation applies to the threshold DM and the inclusive DM.

Item weights are potentially more attractive as they allow experts to up- or down-weight themselves for individual items according to how much they feel they know about that item. "Knowing less" means choosing quantiles further apart and lowering the information score for that item. Of course, the “good performance” of item weights requires that experts can perform this up/down-weighting successfully. Anecdotal evidence suggests that item weights improve over global weights as the experts receive more training in probabilistic assessment. For both global and item weights, calibration dominates over information; information serves to modulate between equally well calibrated experts. Definitions and proofs of these scoring rule properties are found in Cooke (1991) (7).

Optimizing the weights in (2.3) and (2.6) often causes experts to be unweighted, even those with good scores. Such experts are not “rejected”; unweighting simply means that their input is already captured by a smaller subset of experts. Their value to the whole study is brought out in studying the robustness of the optimal DM under the loss of experts and in determining the intrinsic range.

The weights discussed above are all ‘performance based’ (i.e., an expert’s weight depends on their performance). Another performance-based weight is the ‘point-predictive-performance’ weight, or simply ‘predictive-performance’ weight*.* Each item, i, with observed value, O_i_, is divided by the prediction, P_e,i_, of the expert, e, to form the ratio, R_e,i_ = O_i_ / P_e,i_, assuming P_e,i_ > 0*.* For each expert, the exponentiated mean and standard deviation of the logged values of R_e,i_ running over the values of i, termed the geometric mean and geometric standard deviation, are the performance measures for expert. The predictive-performance DM is formed by taking a weighted combination of models’ densities where the weights are proportional to the variance over i of R_e,i_.

It is becoming popular, especially in the climate modeling community, to average the results of models (aka, “experts”). A mathematical justification for this operation is sometimes based on treating model predictions as unbiased estimators with an imputed error term (5). There are many variations of this approach, but the simplest is based on the theory of weighted least squares. Suppose that uncertain quantity, X, for item, i, is estimated by measurement, Z_i_, with error, e_i_. Suppose e_i_ is normally distributed with mean zero and standard deviation σ_i_*.* Then after observing Z_i_ = z_i_, the Renyi conditional distribution of X is normal with mean z_i_ and standard deviation σ_i_. For N models with independent unbiased normal errors, the distribution of X conditional on observing Z_1_ = z_1_, ..., Z_N_ = z_N_ is normal with a mean and variance given by:

E (X | Z_1_ = z_1_, ..., Z_N_ = z_N_) = Σ_i=1, ..., N_ z_i_ w_i_

(2.6)

Var ((X | Z_1_ = z_1_, ..., Z_N_ = z_N_) = 1 / (1 / σ^2^_1_ + ..., 1 / σ^2^_N_)

(2.7)

w_i_ = (1 / σ^2^_N_) / (1 / σ^2^_1_ + ..., 1 / σ^2^_N_)

(2.8)

The weights are proportional to the inverse-variance of each error term. Because of Renyi conditionalization the posterior mean and variance apply to the uncertain quantity, X, and not, as in standard statistical treatments, to the maximum likelihood estimator of X (see Cooke and Wielicki, 2018, for a full discussion) (8). The rest the derivations are entirely standard, and this DM is termed the ‘weighted least squares’ DM.

To apply this theory, we must impute a variance to the assessments of the model for each item*.* The quantile assessments of the models are often not symmetric around the medians. When treating the medians as “observations”, the resulting errors cannot be normal. A somewhat better result is obtained by assuming that the errors are independently and lognormally distributed. The above theory then applies mutatis mutandis to the logged observations. **Table S10** shows a number of these DM’s together with the nine individual models for the first period.

Evidence from many expert judgment applications suggests that roughly 75% of experts would be rejected with statistical hypotheses at the 0.05 level (9). In **Table S10**, three of the nine calibration scores exceeded this threshold. Apart from predictive-performance weighting, all the calibration scores of the performance-weighted DMs were acceptable. The item-weighted DMs tended to be more informative than the global-weighted DMs. Also note the differences between the information scores and the precision scores. Precision scores reflect the lack of variation in the ratio of observed to predicted values. An expert whose prediction is consistently 10 times the observed value will be very precise in this sense (i.e., have a low GSD). The information scores, on the other hand, do not depend on the realizations at all. The non-performance-weighted DMs, equal-weight and weighted least squares, tended to have poor calibration, especially the latter. The inverse-variances in the weighted least squares reflect the self-confidence of the experts without reference to the observed values. It is useful to reflect on this low performance because of the currency of inverse-variance weights in classical and Bayesian model ensemble averaging.

The lognormal weighted least squares DM often tracks the experts with the smallest 90% confidence interval(s). If these very narrow confidence bands capture the realizations with the appropriate relative frequency, then these experts, and consequently the weighted least squares DM, would perform very well. However, it is often the case that very narrow confidence bands fail to catch the realization. Since weighted least squares tracks the ‘overconfident’ experts for each calibration variable separately, its statistical performance was worse than any of the models. Indeed, none of the models are overconfident on *all* calibration variables.

4. **Robustness Analysis**

To determine the robustness against the loss of a single expert, each expert is removed one at a time and the DM is re-calculated. **Table S11** provides these results for models during the summer 2020 period and for the GlobalCM05 DM (i.e., the global-weighted DM with the 0.05 threshold which is the CM-weighted ensemble reported in the manuscript). The rightmost column gives the relative information of each model with respect to the equal-weighted DM, and thus reflects the disagreement among the models themselves. The second column from the right gives the relative information of the perturbed GlobalCM05 DM relative to the unperturbed GlobalCM05 DM*.* When these latter values are much smaller than those in the rightmost column, the perturbation caused by losing a model was small relative to the disagreement among the models themselves. Had we used the GlobalCMOp, or the optimized global-weighted DM, all the weight would have gone to model D and losing this model would produce a perturbation of 0.5, on the same order as the values in the rightmost column. This happens because the best performing model (model D) was rather unlike the other good performers (models A and F). GlobalCM05 was more robust against the loss of a model than GlobalCMOp and although GlobalCM05 exhibited a modest loss in informativeness relative to GlobalCMOp, it still had very good performance. For these reasons the authors chose to adopt the GlobalCM05 DM as the CM-weighted ensemble in the manuscript.

Robustness on items proceeds in the same way. Each calibration item is removed one at a time and the DM is re-calculated. **Table S12** gives results for the summer 2020 models and for GlobalCM05. The relative information with respect to the unperturbed DM, GlobalCM05, is shown in the rightmost column. As with the robustness of the models, the perturbation wrought by losing a single item is small relative to the disagreement among the models themselves.

The winter 2021 performance scores and robustness results are like those of the first period and are not shown.

5. **Determining State Case Rates and Racial Composition; Selecting States**

The COVID-19 case rate for the summer 2020 period was estimated by dividing the number of cases within the week ending August 9^th^ by the state population, and then multiplying by 100,000. Data on cases were taken from the Centers for Disease Control and Prevention (CDC) reported United States cases by state over time (10). State population data came from one-year estimates from the Census Bureau’s 2018 American Community Survey (ACS) (11). The winter 2021 dataset utilized case rates based on cases within the week that ended on March 7^th^ and state population data from one-year estimates provided by the Census Bureau’s 2019 ACS (10, 11).

The fraction of each state’s population reported as non-Hispanic Black and non-Hispanic White was also estimated from the 2018 and 2019 ACS (11). Population data was restricted to the civilian, non-institutionalized population for whom the ACS collects and reports poverty information (11).

Using these data, two states (Louisiana and Idaho for the summer 2020 period; Vermont and Georgia for the winter 2021 period) with case rates in the top 1/3^rd^ of all states, and two states (Maine and New York for the summer; Wyoming and Maryland for the winter) with case rates in the bottom 1/3^rd^ of all states, were selected.

Similarly, two states (Louisiana and New York for the summer; Georgia and Maryland for the winter) in the top 1/3^rd^ of all states in terms of the fraction of their population reported as non-Hispanic Black, and two states (Maine and Idaho for the summer; Vermont and Wyoming for the winter) in the top 1/3^rd^ of all states in terms of the fraction of their population reported as non-Hispanic White, were selected. To select the states that fit these requirements, states with the lowest average rank combination were selected (rank was reversed for the bottom-third case rate requirement).

For the first sensitivity analysis, two states (Kentucky and Maryland for the summer; New Hampshire and Mississippi for the winter) with case rates in the middle 1/3rd of all states were selected. One state (Maryland for the summer; Mississippi for the winter) was in the top 1/3^rd^ of all states in terms of the fraction of their population reported as non-Hispanic Black, and one state (Kentucky for the summer; New Hampshire for the winter) was in the top 1/3^rd^ of all states in terms of the fraction of their population reported as non-Hispanic White.

For the second sensitivity analysis, three states (Oklahoma, Massachusetts, and Rhode Island for the summer; Oklahoma, Rhode Island, and Kansas for the winter) with case rates in the middle 1/3^rd^ of all states were selected. All three of these states were in the middle 1/3^rd^ of all states in terms of fraction of their population reported as non-Hispanic Black, as well as non-Hispanic White.

**Supplemental Figures & Tables**

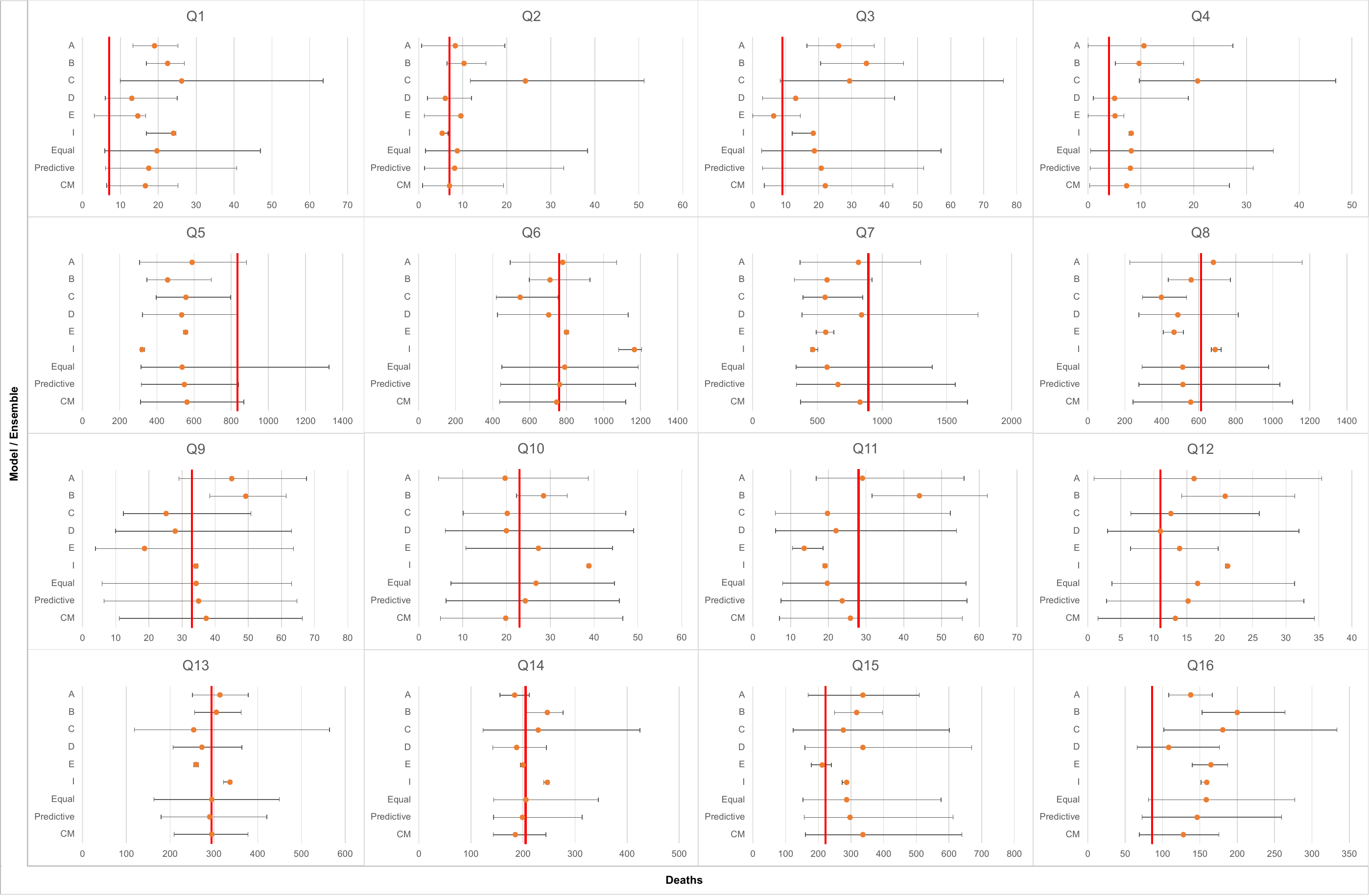

***Abbreviations:*** CM = Classical Model; Q = (week in) question

**Fig. S1:** A forest plot of forecasts provided by the individual models and constructed ensembles for each of the variables of interest for winter 2021. The horizontal axis represents deaths during the week in question, the true values are given by the red vertical lines, the error bars indicate the 5^th^ and 95^th^ percentiles, and the dots represent the model’s predictions.

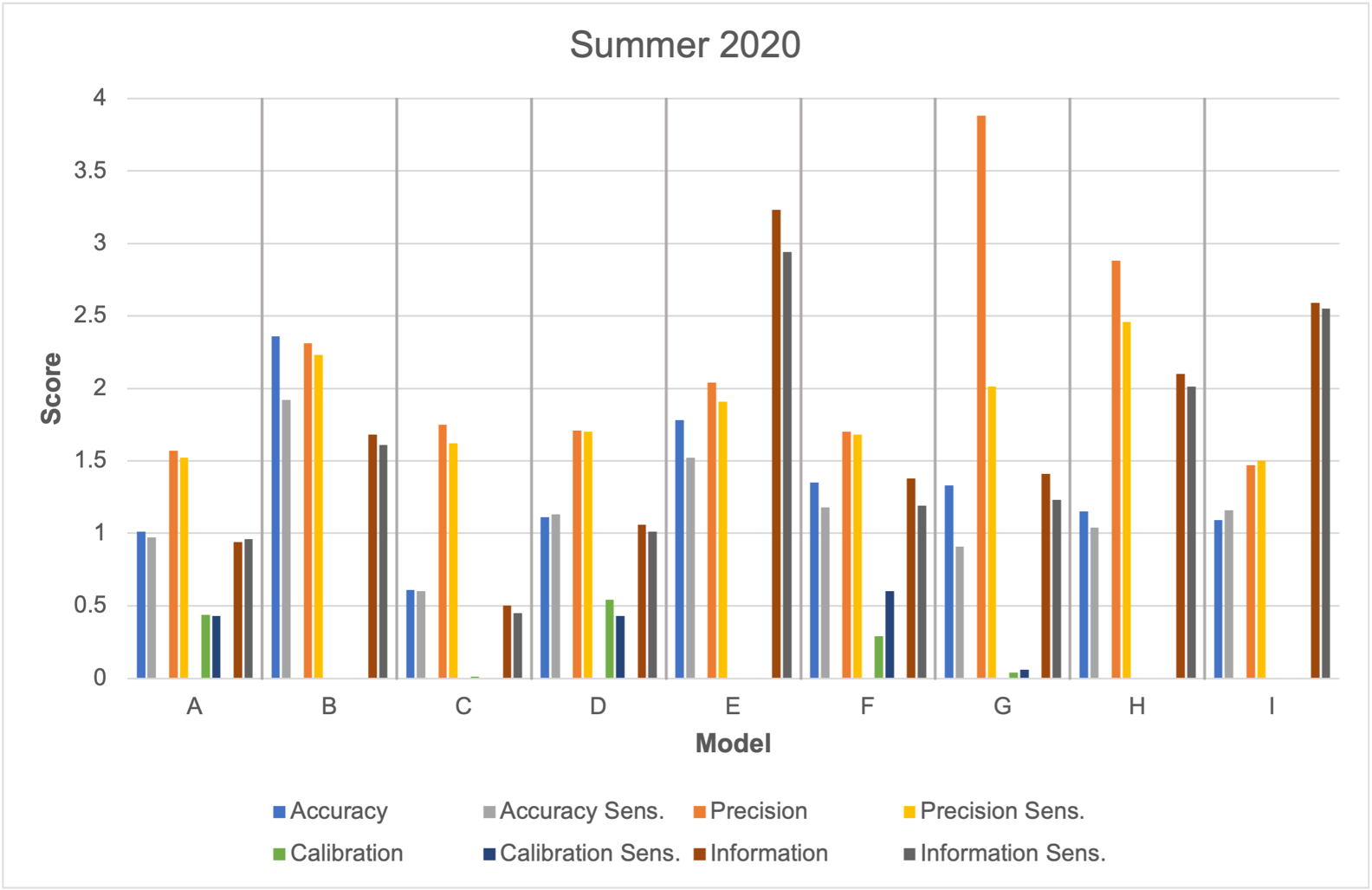

***Note:*** “Sens.” refers to the sensitivity analysis (e.g., “Accuracy Sens.” refers to accuracy in the sensitivity analysis; not, sensitivity of the accuracy parameter).

**Fig. S2:** Performance results of individual model forecasts for summer 2020 that were included in the original dataset and the first sensitivity analysis, which added forecasts for two states (i.e., Kentucky and Maryland) with middling case rates to the original dataset. Shown here are all four performance metric scores for these two datasets for the summer 2020 period.

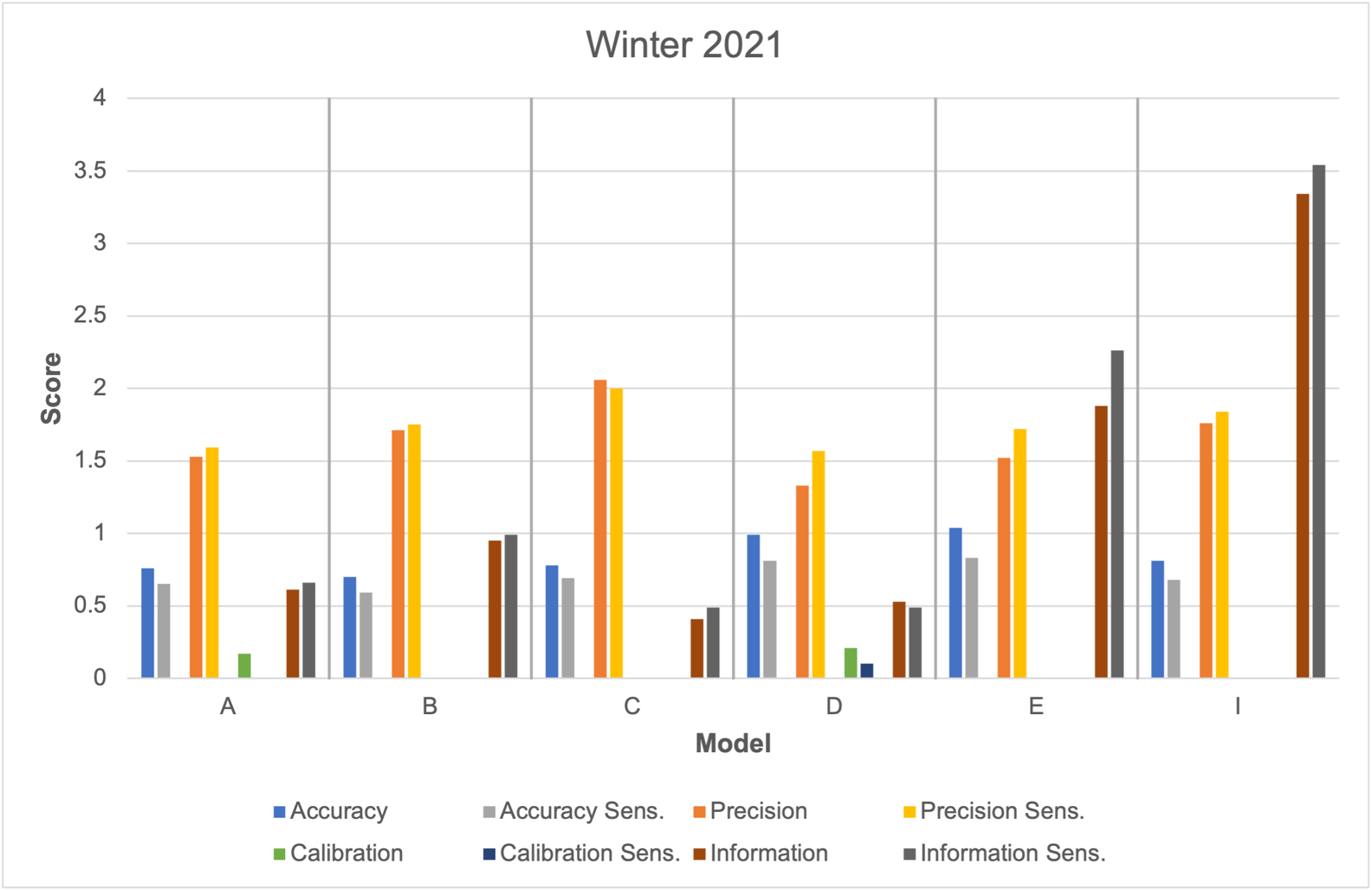

***Note:*** “Sens.” refers to the sensitivity analysis (e.g., “Accuracy Sens.” refers to accuracy in the sensitivity analysis; not, sensitivity of the accuracy parameter).

**Fig. S3:** Performance results of individual model forecasts for winter 2021 that were included in the original dataset and the first sensitivity analysis, which added forecasts for two states (i.e., New Hampshire and Mississippi) with middling case rates to the original dataset. Shown here are all four performance metric scores for these two datasets for the winter 2021 period.

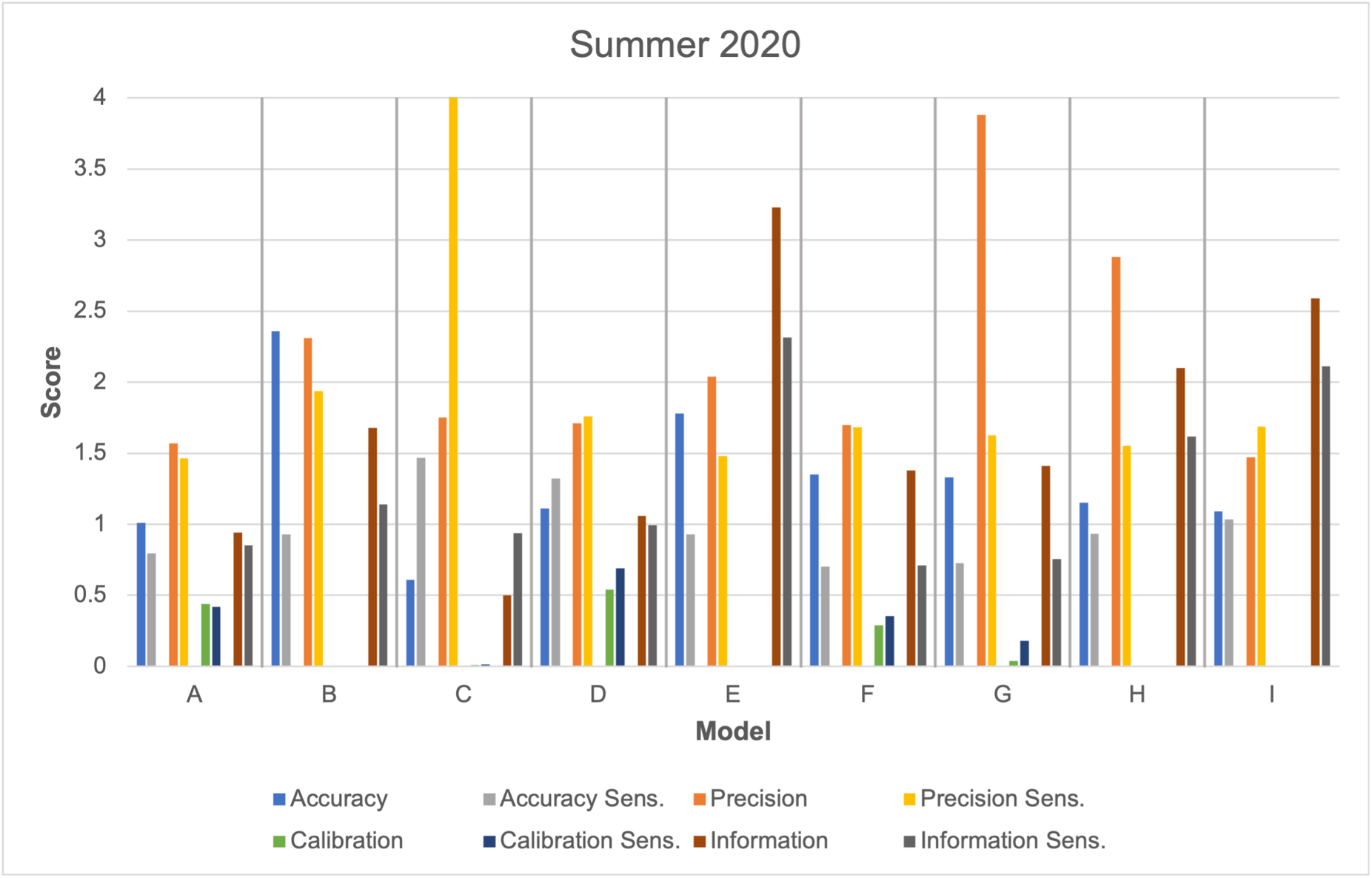

***Note:*** “Sens.” refers to the sensitivity analysis (e.g., “Accuracy Sens.” refers to accuracy in the sensitivity analysis; not, sensitivity of the accuracy parameter).

**Fig. S4:** Performance results of individual model forecasts for summer 2020 that were included in the original dataset and the second sensitivity analysis, which assessed forecasts for three states (i.e., Oklahoma, Massachusetts, Rhode Island) with middling case rates only. Shown here are all four performance metric scores for these two datasets for the summer 2020 period.

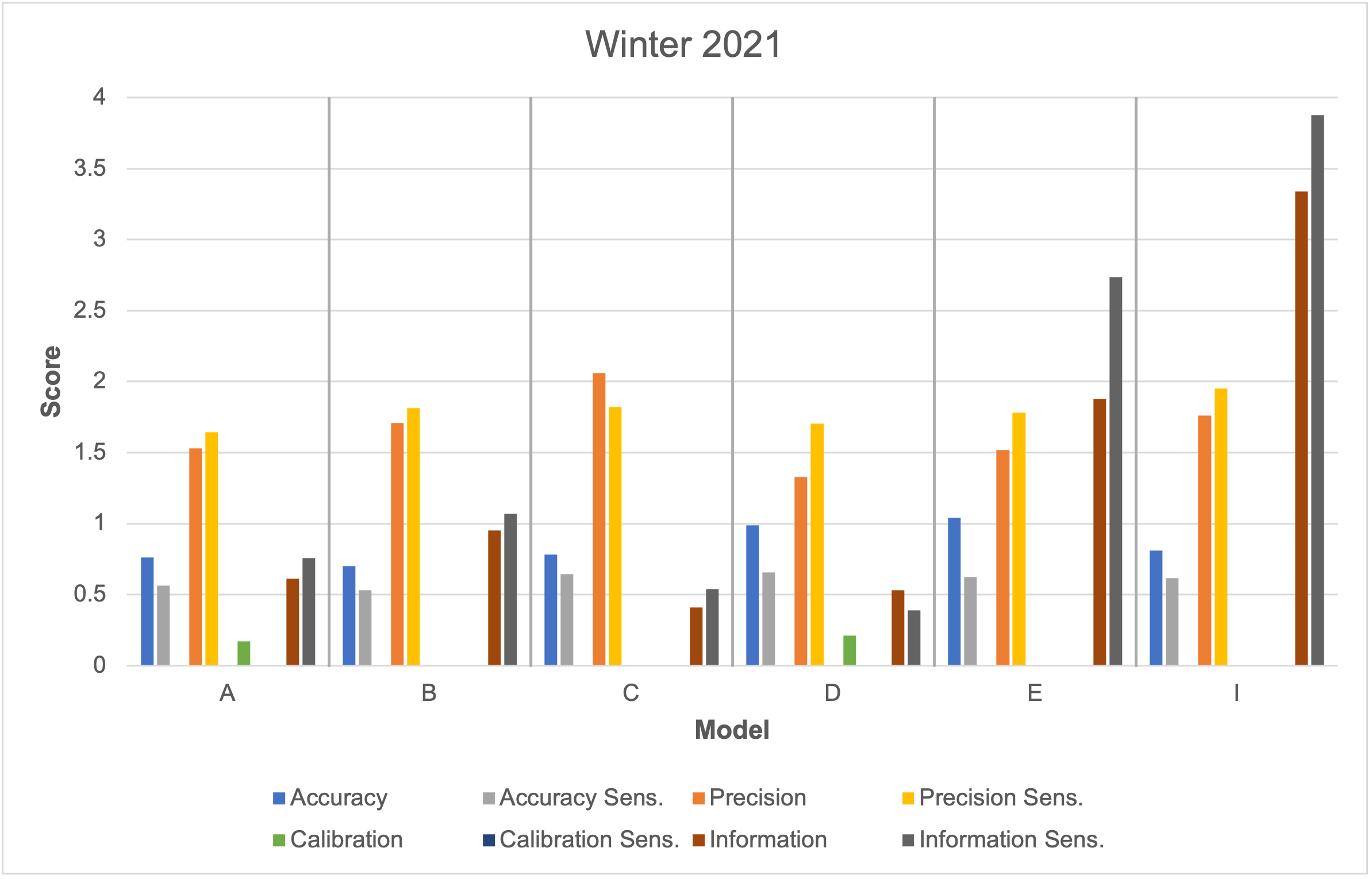

***Note:*** “Sens.” refers to the sensitivity analysis (e.g., “Accuracy Sens.” refers to accuracy in the sensitivity analysis; not, sensitivity of the accuracy parameter).

**Fig. S5:** Performance results of individual model forecasts for winter 2021 that were included in the original dataset and the second sensitivity analysis, which assessed forecasts for three states (i.e., Kansas, Rhode Island, Oklahoma) with middling case rates only. Shown here are all four performance metric scores for these two datasets for the winter 2021 period.

**Table** **S1:** **Individual model performance for summer 2020 (*Left*) and winter 2021 (*Right*)**

| **Model** | **Summer 2020** | | | | |  | **Winter 2021** | | | | |
| --- | --- | --- | --- | --- | --- | --- | --- | --- | --- | --- | --- |
|  | **Accuracy - GM** | **Precision - GSD** | **Calibration** | **Information** | **CM Weight (Unnormalized)** |  | **Accuracy - GM** | **Precision - GSD** | **Calibration** | **Information** | **CM Weight (Unnormalized)** |
| **A** | 1.01 | 1.57 | 0.44 | 0.94 | 0.42 |  | 0.76 | 1.53 | 0.17 | 0.61 | 0.10 |
| **B** | 2.36 | 2.31 | << 0.01 | 1.68 | << 0.01 |  | 0.70 | 1.71 | << 0.01 | 0.95 | << 0.01 |
| **C** | 0.61 | 1.75 | 0.01 | 0.50 | << 0.01 |  | 0.78 | 2.06 | << 0.01 | 0.41 | << 0.01 |
| **D** | 1.11 | 1.71 | 0.54 | 1.06 | 0.57 |  | 0.99 | 1.33 | 0.21 | 0.53 | 0.11 |
| **E** | 1.78 | 2.04 | << 0.01 | 3.23 | << 0.01 |  | 1.04 | 1.52 | 0.01 | 1.88 | 0.03 |
| **F** | 1.35 | 1.70 | 0.29 | 1.38 | 0.40 |  | n/a | n/a | n/a | n/a | n/a |
| **G** | 1.33* | 3.88* | 0.04 | 1.41 | 0.06 |  | n/a | n/a | n/a | n/a | n/a |
| **H** | 1.15 | 2.88 | << 0.01 | 2.10 | << 0.01 |  | n/a | n/a | n/a | n/a | n/a |
| **I** | 1.09 | 1.47 | << 0.01 | 2.59 | << 0.01 |  | 0.81 | 1.76 | << 0.01 | 3.34 | << 0.01 |

******* This value relies on treating model G’s four-weeks ahead prediction of 0 for the week ending on July 11th in Idaho as 0.1. If instead this model G prediction is dropped from the analysis, then the GM becomes 1.00 and the GSD becomes 2.19.

***Abbreviations:*** CM = Classical Model; GM = geometric mean; GSD = geometric standard deviation

**Table S2: Ensemble model performance for summer 2020 using only data from the six models that were available for winter 2021**

| **Ensemble** | **Summer 2020** | | | | |
| --- | --- | --- | --- | --- | --- |
|  | **Accuracy - GM** | **Precision - GSD** | **Calibration** | **Information** | **CM Weight (Unnormalized)** |
| **Equal Weight** | 1.23 | 1.44 | 0.01 | 0.70 | 0.01 |
| **Performance – Predictive Weight** | 0.94 | 1.41 | 0.03 | 0.67 | 0.02 |
| **Performance – CM Weight*** | 1.02 | 1.56 | 0.20 | 0.77 | 0.15 |

***** A hypothesis rejection significance level of 0.05 was used for this ensemble. More detail on why this was used is available in ***SI appendix***, **Notes 3 and 4**.

***Abbreviations:*** CM = Classical Model; GM = geometric mean; GSD = geometric standard deviation

**Table S3: Individual state data on racial composition, coronavirus disease 2019 (COVID-19) cases, total population, and estimated COVID-19 case rate for summer 2020**

| **Location** | **% Non-Hispanic White** | **% Non-Hispanic Black** | **COVID-19 Cases** | **Population** | **COVID-19 Case Rate** |
| --- | --- | --- | --- | --- | --- |
| **Alabama** | 65.3 | 26.6 | 9890 | 4887871 | 202.34 |
| **Alaska** | 60.1 | 3.3 | 431 | 737438 | 58.45 |
| **Arizona** | 54.3 | 4.3 | 8456 | 7171646 | 117.91 |
| **Arkansas** | 72.1 | 15.1 | 5573 | 3013825 | 184.91 |
| **California** | 36.6 | 5.5 | 44998 | 39557045 | 113.75 |
| **Colorado** | 67.8 | 3.9 | 2944 | 5695564 | 51.69 |
| **Connecticut** | 66.3 | 10 | 510 | 3572665 | 14.28 |
| **Delaware** | 61.8 | 21.8 | 685 | 967171 | 70.83 |
| **District of Columbia** | 36.9 | 44.4 | 479 | 702455 | 68.19 |
| **Florida** | 53.3 | 15.3 | 45368 | 21299325 | 213.00 |
| **Georgia** | 52.2 | 31.2 | 24323 | 10519475 | 231.22 |
| **Hawaii** | 21.7 | 1.9 | 1184 | 1420491 | 83.35 |
| **Idaho** | 81.8 | 0.6 | 3327 | 1754208 | 189.66 |
| **Illinois** | 60.9 | 13.8 | 12156 | 12741080 | 95.41 |
| **Indiana** | 78.7 | 9.4 | 6471 | 6691878 | 96.70 |
| **Iowa** | 85.4 | 3.6 | 3297 | 3156145 | 104.46 |
| **Kansas** | 75.6 | 5.6 | 2826 | 2911510 | 97.06 |
| **Kentucky** | 84.5 | 7.8 | 3797 | 4468402 | 84.97 |
| **Louisiana** | 58.4 | 32.2 | 11652 | 4659978 | 250.04 |
| **Maine** | 93 | 1.4 | 81 | 1338404 | 6.05 |
| **Maryland** | 50.2 | 29.5 | 5114 | 6042718 | 84.63 |
| **Massachusetts** | 70.7 | 7 | 2582 | 6902149 | 37.41 |
| **Michigan** | 74.8 | 13.6 | 4965 | 9995915 | 49.67 |
| **Minnesota** | 79.4 | 6.5 | 5569 | 5611179 | 99.25 |
| **Mississippi** | 56.4 | 37.8 | 6048 | 2986530 | 202.51 |
| **Missouri** | 79.3 | 11.4 | 7087 | 6126452 | 115.68 |
| **Montana** | 85.9 | 0.5 | 824 | 1062305 | 77.57 |
| **Nebraska** | 78.5 | 4.6 | 1730 | 1929268 | 89.67 |
| **Nevada** | 48.4 | 8.8 | 6019 | 3034392 | 198.36 |
| **New Hampshire** | 89.8 | 1.4 | 197 | 1356458 | 14.52 |
| **New Jersey** | 54.6 | 12.8 | 2423 | 8908520 | 27.20 |
| **New Mexico** | 36.9 | 1.9 | 1299 | 2095428 | 61.99 |
| **New York** | 55.2 | 14.3 | 2453 | 19542209 | 12.55 |
| **North Carolina** | 62.7 | 21 | 11511 | 10383620 | 110.86 |
| **North Dakota** | 83.8 | 3.4 | 928 | 760077 | 122.09 |
| **Ohio** | 78.6 | 12.2 | 7817 | 11689442 | 66.87 |
| **Oklahoma** | 65.2 | 7.2 | 5742 | 3943079 | 145.62 |
| **Oregon** | 75.1 | 1.9 | 2175 | 4190713 | 51.90 |
| **Pennsylvania** | 75.9 | 10.6 | 5262 | 12807060 | 41.09 |
| **Rhode Island** | 71.4 | 5.8 | 716 | 1057315 | 67.72 |
| **South Carolina** | 63.5 | 26.5 | 8647 | 5084127 | 170.08 |
| **South Dakota** | 81.5 | 2.1 | 650 | 882235 | 73.68 |
| **Tennessee** | 73.6 | 16.6 | 13085 | 6770010 | 193.28 |
| **Texas** | 41.4 | 11.9 | 55877 | 28701845 | 194.68 |
| **Utah** | 77.8 | 1.2 | 2955 | 3161105 | 93.48 |
| **Vermont** | 92.6 | 1.2 | 36 | 626299 | 5.75 |
| **Virginia** | 61.3 | 18.8 | 7644 | 8517685 | 89.74 |
| **Washington** | 67.8 | 3.7 | 4900 | 7535591 | 65.02 |
| **West Virginia** | 92 | 3.8 | 840 | 1805832 | 46.52 |
| **Wisconsin** | 81 | 6.3 | 5845 | 5813568 | 100.54 |
| **Wyoming** | 84 | 0.5 | 242 | 577737 | 41.89 |

**Table S4: Individual state data on racial composition, coronavirus disease 2019 (COVID-19) cases, total population, and estimated COVID-19 case rate for winter 2021**

| **Location** | **% Non-Hispanic White** | **% Non-Hispanic Black** | **COVID-19 Cases** | **Population** | **COVID-19 Case Rate** |
| --- | --- | --- | --- | --- | --- |
| **Alabama** | 65.1 | 26.8 | 6567 | 4903185 | 133.93 |
| **Alaska** | 59.8 | 3 | 897 | 731545 | 122.62 |
| **Arizona** | 54 | 4.4 | 9672 | 7278717 | 132.88 |
| **Arkansas** | 72 | 15.4 | 2403 | 3017804 | 79.63 |
| **California** | 36.3 | 5.5 | 25832 | 39512223 | 65.38 |
| **Colorado** | 67.5 | 4 | 8299 | 5758736 | 144.11 |
| **Connecticut** | 65.6 | 10.1 | 5384 | 3565287 | 151.01 |
| **Delaware** | 61.3 | 22 | 1555 | 973764 | 159.69 |
| **District of Columbia** | 37.3 | 44.1 | 821 | 705749 | 116.33 |
| **Florida** | 53 | 15.2 | 35055 | 21477737 | 163.22 |
| **Georgia** | 51.8 | 31.5 | 16966 | 10617423 | 159.79 |
| **Hawaii** | 21.5 | 1.8 | 310 | 1415872 | 21.89 |
| **Idaho** | 81.6 | 0.7 | 1791 | 1787065 | 100.22 |
| **Illinois** | 60.7 | 13.9 | 11639 | 12671821 | 91.85 |
| **Indiana** | 78.3 | 9.4 | 5589 | 6732219 | 83.02 |
| **Iowa** | 85.1 | 4 | 3245 | 3155070 | 102.85 |
| **Kansas** | 75.4 | 5.5 | 2198 | 2913314 | 75.45 |
| **Kentucky** | 84.2 | 8 | 6087 | 4467673 | 136.25 |
| **Louisiana** | 58.2 | 32.1 | 3685 | 4648794 | 79.27 |
| **Maine** | 92.8 | 1.5 | 1163 | 1344212 | 86.52 |
| **Maryland** | 49.8 | 29.7 | 5333 | 6045680 | 88.21 |
| **Massachusetts** | 70.3 | 7.1 | 8781 | 6892503 | 127.40 |
| **Michigan** | 74.7 | 13.5 | 10522 | 9986857 | 105.36 |
| **Minnesota** | 78.9 | 6.4 | 5417 | 5639632 | 96.05 |
| **Mississippi** | 56.3 | 37.8 | 2657 | 2976149 | 89.28 |
| **Missouri** | 79.1 | 11.4 | 4296 | 6137428 | 70.00 |
| **Montana** | 85.8 | 0.6 | 960 | 1068778 | 89.82 |
| **Nebraska** | 78.4 | 4.8 | 2145 | 1934408 | 110.89 |
| **Nevada** | 47.8 | 9.3 | 2436 | 3080156 | 79.09 |
| **New Hampshire** | 89.7 | 1.4 | 1437 | 1359711 | 105.68 |
| **New Jersey** | 54.3 | 12.7 | 23253 | 8882190 | 261.79 |
| **New Mexico** | 36.8 | 1.9 | 1790 | 2096829 | 85.37 |
| **New York** | 55.1 | 14.2 | 23322 | 19453561 | 119.89 |
| **North Carolina** | 62.5 | 21.1 | 15130 | 10488084 | 144.26 |
| **North Dakota** | 83.6 | 2.9 | 567 | 762062 | 74.40 |
| **Ohio** | 78.3 | 12.4 | 11049 | 11689100 | 94.52 |
| **Oklahoma** | 64.9 | 7.1 | 3852 | 3956971 | 97.35 |
| **Oregon** | 74.9 | 1.8 | 1688 | 4217737 | 40.02 |
| **Pennsylvania** | 75.6 | 10.7 | 17001 | 12801989 | 132.80 |
| **Rhode Island** | 70.8 | 5.8 | 2499 | 1059361 | 235.90 |
| **South Carolina** | 63.5 | 26.3 | 7656 | 5148714 | 148.70 |
| **South Dakota** | 81.5 | 2.3 | 1162 | 884659 | 131.35 |
| **Tennessee** | 73.3 | 16.6 | 8480 | 6829174 | 124.17 |
| **Texas** | 41.1 | 11.9 | 42794 | 28995881 | 147.59 |
| **Utah** | 77.7 | 1.1 | 3615 | 3205958 | 112.76 |
| **Vermont** | 92.5 | 1.3 | 885 | 623989 | 141.83 |
| **Virginia** | 61.1 | 19 | 9418 | 8535519 | 110.34 |
| **Washington** | 67.3 | 3.9 | 5039 | 7614893 | 66.17 |
| **West Virginia** | 92 | 3.6 | 1590 | 1792147 | 88.72 |
| **Wisconsin** | 80.8 | 6.3 | 4287 | 5822434 | 73.63 |
| **Wyoming** | 83.7 | 1.1 | 370 | 578759 | 63.93 |

**Table S5: Individual model and constructed ensemble forecasts of coronavirus disease 2019 (COVID-19) mortality for summer 2020 stratified by quantile, forecast timeframe, and state, with subsequent observations**

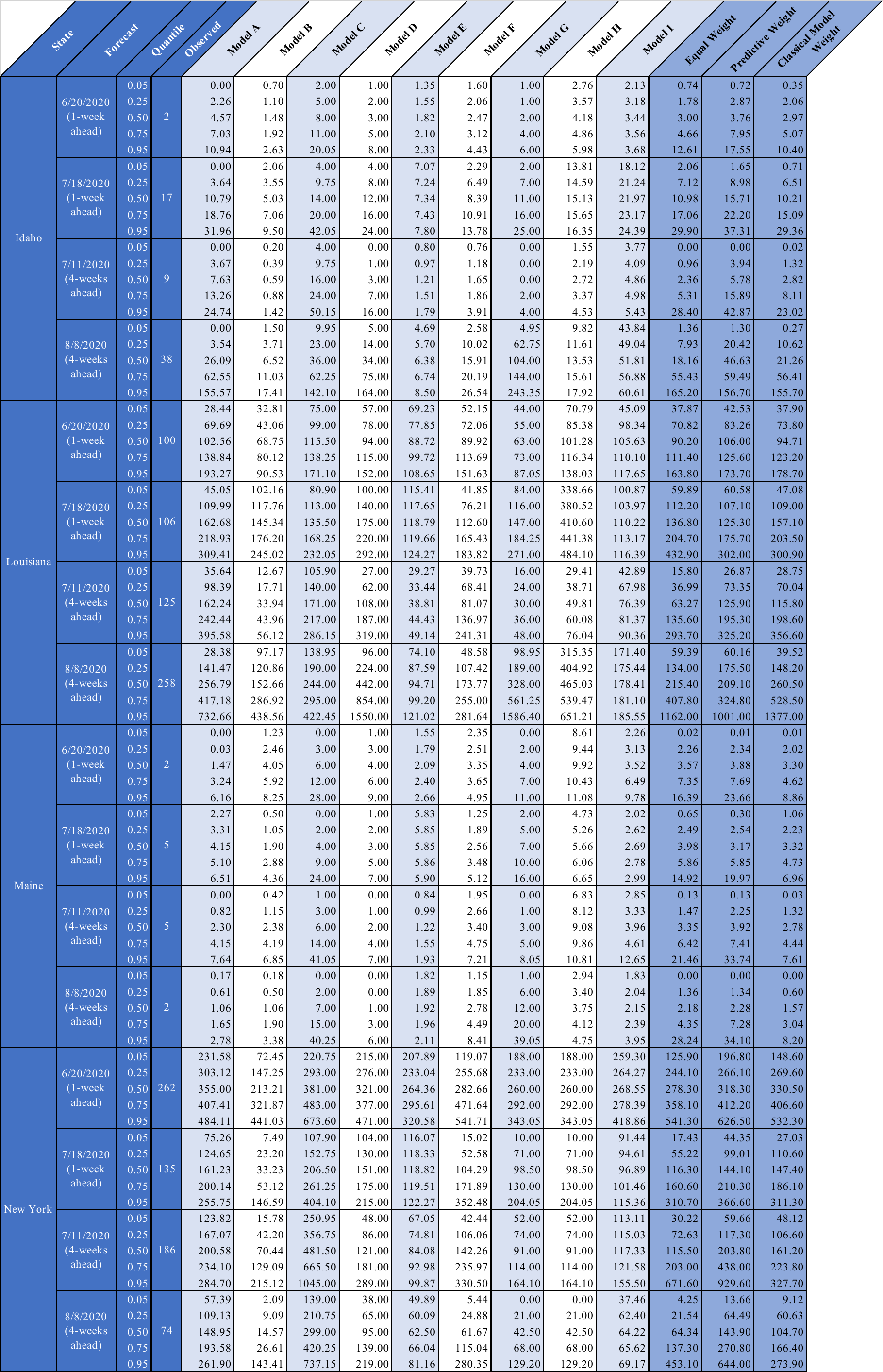

**Table S6: Individual model and constructed ensemble forecasts of coronavirus disease 2019 (COVID-19) mortality for summer 2020 stratified by quantile, forecast timeframe, and state, with subsequent observations**

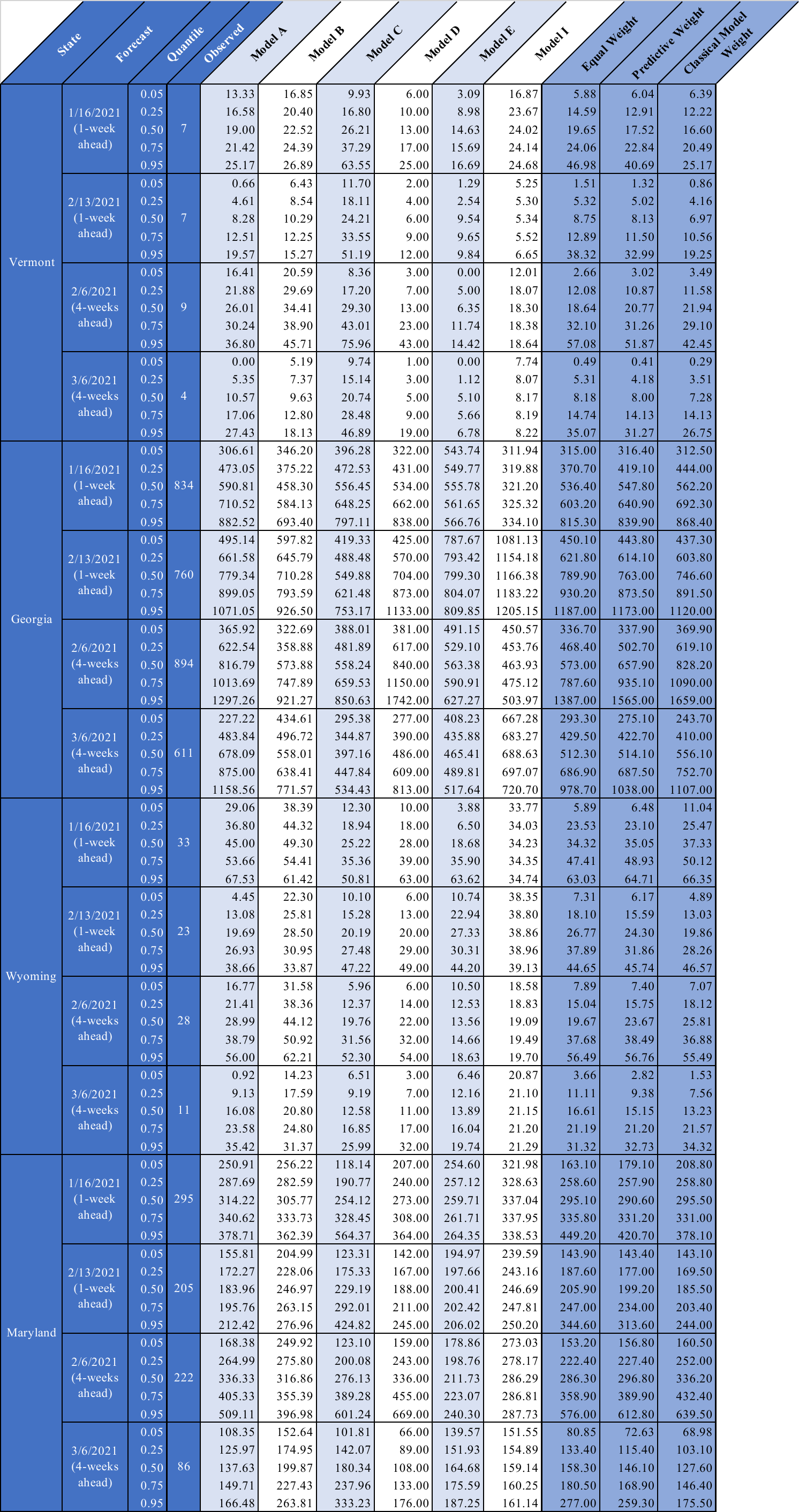

**Table S7: Classical-Model-weighted ensemble model performance for summer 2020 (*Left*) and winter 2021 (*Right*) stratified by different domains**

| **Sub-Domain** | **Summer 2020** | | | | |  | **Winter 2021** | | | | |
| --- | --- | --- | --- | --- | --- | --- | --- | --- | --- | --- | --- |
|  | **Accuracy - GM** | **Precision – GSD** | **Calibration** | **Information** | **CM Weight (Unnormalized)** |  | **Accuracy - GM** | **Precision – GSD** | **Calibration** | **Information** | **CM Weight (Unnormalized)** |
| **High % non-Hispanic White**  **(ID, ME; VT, WY)** | 1.38 | 1.73 | 0.14 | 1.08 | 0.16 |  | 0.74 | 1.52 | 0.33 | 0.46 | 0.15 |
| **High % non-Hispanic Black**  **(LA, NY; GA, MD)** | 0.91 | 1.22 | 0.33 | 0.54 | 0.18 |  | 0.98 | 1.31 | 0.73 | 0.40 | 0.29 |
| **High Case Rate**  **(ID, LA; VT, GA)** | 1.22 | 1.70 | 0.73 | 0.57 | 0.41 |  | 0.80 | 1.63 | 0.53 | 0.45 | 0.24 |
| **Low Case Rate**  **(ME, NY; WY. MD)** | 1.03 | 1.46 | 0.88 | 1.05 | 0.92 |  | 0.91 | 1.25 | 0.79 | 0.41 | 0.33 |
| **One Week Ahead** | 0.92 | 1.46 | 0.36 | 0.63 | 0.23 |  | 0.96 | 1.44 | 0.79 | 0.52 | 0.41 |
| **Four Weeks Ahead** | 1.36 | 1.59 | 0.27 | 0.98 | 0.26 |  | 0.76 | 1.44 | 0.29 | 0.34 | 0.10 |

***Abbreviations:*** CM = Classical Model; GM = geometric mean; GSD = geometric standard deviation; GA = Georgia; ID = Idaho; LA = Louisiana; MD = Maryland; ME = Maine; NY = New York; VT = Vermont; WY = Wyoming

**Table S8: Predictive-performance-weighted ensemble model performance for summer 2020 (*Left*) and winter 2021 (*Right*) stratified by different domains**

| **Sub-Domain** | **Summer 2020** | | | | |  | **Winter 2021** | | | | |
| --- | --- | --- | --- | --- | --- | --- | --- | --- | --- | --- | --- |
|  | **Accuracy - GM** | **Precision – GSD** | **Calibration** | **Information** | **CM Weight (Unnormalized)** |  | **Accuracy - GM** | **Precision – GSD** | **Calibration** | **Information** | **CM Weight (Unnormalized)** |
| **High % non-Hispanic White**  **(ID, ME; VT, WY)** | 0.95 | 1.55 | 0.27 | 0.63 | 0.17 |  | 0.70 | 1.50 | 0.16 | 0.36 | 0.06 |
| **High % non-Hispanic Black**  **(LA, NY; GA, MD)** | 0.88 | 1.28 | 0.12 | 0.47 | 0.06 |  | 1.02 | 1.36 | 0.53 | 0.34 | 0.18 |
| **High Case Rate**  **(ID, LA; VT, GA)** | 0.96 | 1.37 | 0.33 | 0.52 | 0.17 |  | 0.81 | 1.70 | 0.73 | 0.39 | 0.29 |
| **Low Case Rate**  **(ME, NY; WY. MD)** | 0.87 | 1.48 | 0.27 | 0.58 | 0.15 |  | 0.88 | 1.26 | 0.33 | 0.30 | 0.10 |
| **One Week Ahead** | 0.86 | 1.44 | 0.20 | 0.46 | 0.09 |  | 0.91 | 1.36 | 0.60 | 0.38 | 0.23 |
| **Four Weeks Ahead** | 0.98 | 1.40 | 0.04 | 0.64 | 0.03 |  | 0.78 | 1.54 | 0.14 | 0.32 | 0.04 |

***Abbreviations:*** CM = Classical Model; GM = geometric mean; GSD = geometric standard deviation; GA = Georgia; ID = Idaho; LA = Louisiana; MD = Maryland; ME = Maine; NY = New York; VT = Vermont; WY = Wyoming

Table S9: Test of random expert hypothesis based on 1000 scrambled panels

|  | Original Panel | | | Random Scrambles | | |
| --- | --- | --- | --- | --- | --- | --- |
| Summer |  |  |  |  |  |  |
|  | Average | Max | Min | %<Av | %<Max | %>Min |
| Calibration | 0.10 | 0.34 | 7.34E-10 | 62 | 41 | 100 |
| Information | 1.55 | 3.12 | 0.43 | NA | 100 | 100 |
| Winter |  | | |  | | |
|  | Average | Max | Min | %<Av | %<Max | %>Min |
| Calibration | 0.05 | 0.22 | 8.79E-17 | 81 | 74 | 100 |
| Information | 1.22 | 3.15 | 0.36 | NA | 100 | 100 |

***Abbreviations:*** %<Av = percentage of scrambled panels in which the average calibration/information/combined was less than the panel average; %<Max = percentage of scrambled panels in which the max calibration/information/combined was less than the panel maximum; %>Min = percentage of scrambled panels in which the minimum calibration/information/combined was greater than the panel minimum

***Note:*** All results were based on 5%, 50%, 95% values; 25% and 75% values were not used.

Table S10: Comparison of model and ensemble performance for summer 2020

|  |  | Assessor | Calibration | Information | CM Weight (Unnormalized) | GlobalCM05 Weight (Normalized) | Accuracy | Precision | Predictive Weights (Unnormalized) | Predictive Weights (Normalized) |
| --- | --- | --- | --- | --- | --- | --- | --- | --- | --- | --- |
| Models | A | OliverWy | 0.44 | 0.94 | 0.42 | **0.30** | 1.01 | 1.57 | 4.19 | 0.18 |
|  | B | MOBS | 5.53E-05 | 1.68 | 0.00 | **0** | 2.36 | 2.31 | 0.08 | 0.00 |
|  | C | JHU | 0.01 | 0.50 | 0.00 | **0** | 0.61 | 1.75 | 8.79 | 0.39 |
|  | D | UMass | 0.54 | 1.06 | 0.57 | **0.41** | 1.11 | 1.71 | 1.92 | 0.08 |
|  | E | UCLA | 1.89E-06 | 3.23 | 0.00 | **0** | 1.78 | 2.04 | 0.26 | 0.01 |
|  | F | YYG | 0.29 | 1.38 | 0.40 | **0.29** | 1.35 | 1.70 | 0.76 | 0.03 |
|  | G | UT | 0.04 | 1.41 | 0.06 | **0** | 1.33 | 3.88 | 1.06 | 0.05 |
|  | H | ERDC | 6.62E-09 | 2.10 | 0.00 | **0** | 1.15 | 2.88 | 0.21 | 0.01 |
|  | I | Covid19Sim | 8.42E-07 | 2.59 | 0.00 | **0** | 1.09 | 1.47 | 5.54 | 0.24 |
| Perf. Ensembles |  | **GlobalCM05** | **0.54** | **0.81** | **0.43** |  | 1.12 | 1.57 |  |  |
|  |  | ItemCM05 | 0.47 | 0.91 | 0.43 |  | 1.20 | 1.70 |  |  |
|  |  | GlobalCMInclusive | 0.54 | 0.77 | 0.42 |  | 1.12 | 1.57 |  |  |
|  |  | ItemCMInclusive | 0.44 | 0.90 | 0.40 |  | 1.22 | 1.70 |  |  |
|  |  | GlobalCMOp^*^ | 0.54 | 1.06 | 0.57 |  | 1.11 | 1.71 |  |  |
|  |  | Predictive | 0.04 | 0.55 | 0.02 |  | 0.91 | 1.41 |  |  |
| Non-Perf. Ensembles |  | Equal | 0.03 | 0.72 | 0.02 |  | 1.19 | 1.55 |  |  |
|  |  | Weighted least squares Log | 4.10E-09 | 3.02 | 1.24E-08 |  | 1.04 | 1.67 |  |  |

^*^GlobalCMOp and ItemCMOp coincide in this case.

***Abbreviations:*** Global = average information over all calibration variables; Item = item specific weights, using the information scores per item; CM = Classical Model; 05 = threshold cutoff at 0.05; Inclusive = no threshold cutoff; Op = optimized

***Note:*** The preferred ensemble for this study is GlobalPw05 (referred to as CM-weighted in the manuscript). For this DM the normalized weights used for combining the models are shown in red.

**Table S11: GlobalCM05 ensemble (i.e., the Classical Model ensemble that averages information over all calibration variables and has a 0.05 threshold)** **robustness against the loss of a single model for summer 2020**

|  | Excluded Model | Information | Calibration | Rel. Information to Orig. CM | Rel. Information to EW Ens. |
| --- | --- | --- | --- | --- | --- |
| A | OliverWy | 0.89 | 0.61 | 0.09 | 0.52 |
| B | MOBS | 0.80 | 0.54 | 0.0005 | 0.62 |
| C | JHU | 0.47 | 0.34 | 0.02 | 0.59 |
| D | UMass | 0.86 | 0.44 | 0.08 | 0.44 |
| E | UCLA | 0.81 | 0.54 | 4.37E-09 | 1.88 |
| F | YYG | 0.81 | 0.20 | 0.10 | 0.47 |
| G | UT | 0.79 | 0.54 | 0.0005 | 0.79 |
| H | ERDC | 0.78 | 0.54 | 0.001 | 1.42 |
| I | Covid19Sim | 0.81 | 0.54 | 1.14E-07 | 1.64 |
|  | GlobalCM05 | 0.81 | 0.54 |  |  |

***Abbreviations:*** Rel. = relative; Orig. = original; CM = Classical Model; EW = equal-weighted; Ens. = ensemble

Table S12: Robustness on items for summer 2020

|  | Excluded Item | Information | Calibration | Rel. Information to Orig. |
| --- | --- | --- | --- | --- |
| 1 | ID 6/20 | 0.80 | 0.53 | 0.06 |
| 2 | ID 7/11 | 0.76 | 0.48 | 0.07 |
| 3 | ID 7/18 | 0.86 | 0.51 | 0.03 |
| 4 | ID 8/8 | 0.80 | 0.57 | 0.07 |
| 5 | LA 6/20 | 0.80 | 0.53 | 0.08 |
| 6 | LA 7/11 | 0.80 | 0.64 | 0.08 |
| 7 | LA 7/18 | 0.78 | 0.32 | 0.06 |
| 8 | LA 8/8 | 0.81 | 0.51 | 0.02 |
| 9 | ME 6/20 | 0.74 | 0.48 | 0.07 |
| 10 | ME 7/11 | 0.77 | 0.57 | 0.02 |
| 11 | ME 7/18 | 0.75 | 0.53 | 0.04 |
| 12 | ME 8/8 | 0.74 | 0.65 | 0.06 |
| 13 | NY 6/20 | 0.82 | 0.54 | 0.02 |
| 14 | NY 7/11 | 0.74 | 0.53 | 0.08 |
| 15 | NY 7/18 | 0.85 | 0.37 | 0.02 |
| 16 | NY 8/8 | 0.80 | 0.42 | 0.01 |
|  | GlobalCM05 | 0.81 | 0.54 |  |

***Abbreviations:*** Rel. = relative; Orig. = original; ID = Idaho; LA = Louisiana; ME = Maine; NY = New York; Global = average information over all calibration variables; CM = Classical Model; 05 = threshold cutoff at 0.05

1. You can see this in the continuous from of the entropy integral. For a discrete distribution *P = p1, ..., pn,* the entropy is defined as *H(P) = -* Σ*pi ln(pi).* To pass to the continuous version replace *Σ → ∫* and *pi → f(x)dx.* That gives *H(f) = -∫ fx(dx) ln(f(x)dx),* which is meaningless. You must use relative information *I (f | g) = ∫ f(x)dx ln(f(x)dx/g(x)dx)* so that the *dx’s* inside the *ln* cancel and *I(f | g) = ∫f(x)dx ln(f(x)/g(x)).*
